# Saturation genome editing of *BRCA1* across cell types accurately resolves cancer risk

**DOI:** 10.1101/2025.08.11.25333423

**Authors:** Phoebe Dace, Nicole M. Forrester, Maria Zanti, Laura Cubitt, Chloé Terwagne, Megan Buckley, Elke M. van Veen, Thomas Stuart Wilson, Paola Scaffidi, Kyriaki Michailidou, Gregory M. Findlay

**Affiliations:** Genome Function Laboratory, The Francis Crick Institute, London, UK; Biostatistics Unit, The Cyprus Institute of Neurology and Genetics, Nicosia, Cyprus; Department of Human Genetics, Research Institute for Medical Innovation, Radboud University Medical Center, Nijmegen, Netherlands; Cancer Epigenetics Laboratory, The Francis Crick Institute, London, UK; Cancer Epigenetics, Department of Experimental Oncology, IEO, European Institute of Oncology IRCCS, Milan, Italy

## Abstract

Germline pathogenic *BRCA1* variants predispose women to breast and ovarian cancer. Despite accumulation of functional evidence for variants in *BRCA1*, over half of reported single-nucleotide variants (SNVs) lack a definitive clinical interpretation. Furthermore, the extent to which variant effects are consistent across cell types remains largely unexplored. Here, we performed saturation genome editing (SGE) of *BRCA1* in HAP1 cells to score 4,113 variants not previously assayed. Additionally, we developed a new SGE assay in human mammary epithelial cells (HMECs), allowing effects of variants to be compared across cell lines, drug treatments, and genetic backgrounds. We identify 538 variants impacting function via diverse mechanisms, including impairment of the BRCA1–PALB2 interaction and disruption of splicing, transcription, and translation. Function scores from experiments in HAP1 discriminate known pathogenic and benign variants with near-perfect accuracy. Intriguingly, however, nearly half of variants impacting function in HAP1 were found to be neutral when assayed in HMECs. We show that discordantly scored variants are hypomorphic and confer intermediate cancer risk. These results will be highly valuable for clinical interpretation of *BRCA1* variants. Moreover, this work illustrates how revealing context-specific variant effects across cell types can enable more accurate resolution of disease risk.

## INTRODUCTION

Clinical interpretation of rare genetic variants remains challenging, even for variants in well-studied genes. This is evidenced by large numbers of variants of uncertain significance (VUS) reported in genes of high clinical importance^1^. Failure to identify variants underlying disease prevents accurate diagnosis and precludes opportunities to improve care.

Functional evidence can inform not only whether a variant is pathogenic or benign, but also the degree of phenotypic effect, which can be of considerable importance for understanding disease penetrance and severity. Recently, multiplexed assays of variant effect (MAVEs) have emerged to functionally interrogate variants at scale^2^. MAVEs enable systematic comparison of thousands of variants per experiment by leveraging next-generation sequencing (NGS)-based readouts^3^. High-quality MAVE data can improve clinical variant classification^4^ and have proven particularly beneficial for classifying variants observed in populations underrepresented in genomics studies^5^.

One MAVE highly suited to distinguishing pathogenic and benign variants is saturation genome editing (SGE)^6^. A key advantage of SGE is that variants are introduced endogenously via CRISPR, such that variants impacting gene function at any level (e.g., splicing, protein folding) can be determined. SGE has performed strongly at distinguishing pathogenic and benign variants when applied to tumour suppressor genes including *BRCA1*, *BRCA2*, *VHL*, *BAP1*, and *RAD51C*^7–15^. A limitation of SGE, and more broadly of MAVEs in general, however, is that most assays are performed only in a single cell type suitable for high-throughput experimentation. To what extent variant effects are consistent across cell types remains largely unknown.

*BRCA1* is a tumour suppressor gene in which pathogenic germline variants greatly increase risk of breast and ovarian cancer in women. Preventative strategies offered to carriers of pathogenic *BRCA1* variants reduce cancer-associated risks^16,17^. Given the importance of identifying *BRCA1* variants of functional consequence, dozens of assays have been deployed to study *BRCA1* variants’ effects^18^, including assays that measure mRNA expression and splicing^6,19,20^, specific BRCA1 functions, such as homologous recombination (HR)^21,22^ and replication fork protection^23^, as well as global effects on cellular fitness^7^.

While MAVE data have been generated for regions of *BRCA1*, no study has systematically assessed the extent to which variant effects are consistent across cell types. This is a critical question not only for *BRCA1* but also many other genes to which MAVEs have been applied, as sufficient weight can be given to functional evidence produced in a single cell type to change how variants are managed clinically.

Here, we perform SGE in human HAP1 cells to test 4,740 *BRCA1* variants and develop an additional SGE assay in human mammary epithelial cells (HMECs) to systematically compare variant effects across cell types. We uncover a substantial fraction of variants with discordant effects across models, ultimately revealing new insights into *BRCA1* function and enabling better estimation of cancer risk.

## RESULTS

### An optimised SGE assay in HAP1 identifies new loss-of-function variants in *BRCA1*

SGE was previously performed in HAP1 to characterise 3,893 SNVs across 13 *BRCA1* exons encoding the RING and BRCT domains.^7^ While most missense variants currently deemed pathogenic in ClinVar map to these domains, it remains unknown to what extent variants elsewhere contribute to disease burden, both in coding and non-coding regions. Despite extensive efforts to profile *BRCA1* variants^18^, as of February 2025, 59.5% of the 6,397 *BRCA1* variants reported outside the RING and BRCT domains are VUS or have conflicting interpretations in ClinVar. (We collectively reference such variants as VUS henceforth.)

We performed SGE experiments in HAP1 cells for 11 regions of *BRCA1* outside the RING and BRCT domains (RefSeq transcript: NM_007294.4; **Fig. 1A**). These included regions covering exons 11 and 12, which encode the coiled-coil (CC) domain, multiple regions of exon 10, exon 6, and non-coding regions where pathogenic variants have been reported, including a deep intronic region of intron 11, the proximal promoter, and the 5’-untranslated region (5’-UTR). Homology-directed DNA repair (HDR) libraries contained between 336 and 402 SNVs, with libraries for the promoter and 5’-UTR also containing tiling 1- and 5-bp deletions.

SGE was performed using a protocol optimised to enable precise variant scoring^12^. For each SGE region, 18 million HAP1 cells were transfected per replicate to deliver a single guide RNA (gRNA) and HDR library. Cells were cultured for 14 days and genomic DNA (gDNA) and RNA were harvested on days 5 and 14. Amplicon sequencing was performed to determine a function score for each variant, corresponding to the log2-ratio of variant frequency on day 14 over day 5 (**Methods**). To reveal variants impacting *BRCA1* mRNA levels, we also performed targeted RNA sequencing of day 5 samples to calculate an RNA score for each coding variant.

In total, 3,843 SNVs and 270 small deletions were assigned function scores across the 11 new SGE regions, including 98.0% of all possible SNVs across these regions (**Fig. 1B, Tables S1, S2**). Function scores were highly reproducible across biological replicates (**Fig. S1A**).

**Figure 1.**
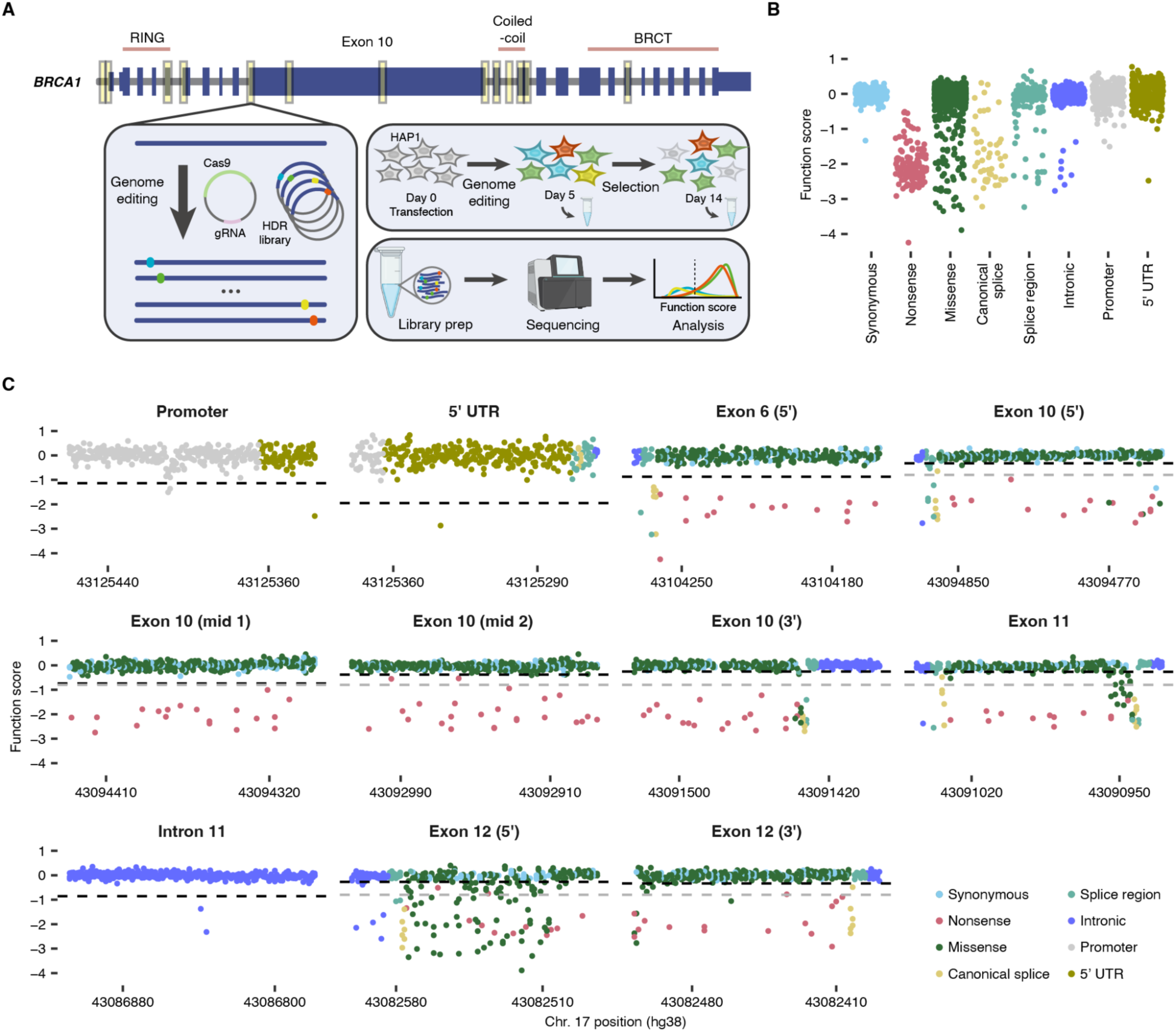
HAP1 SGE reveals new loss-of-function variants in *BRCA1*. **A.** SGE experiments were performed in duplicate for 11 regions of *BRCA1*, including the promoter, non-coding exon 1, coding exons 6, 10, 11, and 12, and intron 11. Exons 5 and 17, in the RING and BRCT domains, respectively, were assayed for comparison to prior data. Yellow boxes indicate SGE regions in this study. HAP1 cells were edited to install an HDR library containing each possible SNV. Function scores, reflecting variants’ effects on fitness, were determined by performing amplicon sequencing to quantify variant frequencies on days 5 and 14. **B.** HAP1 function scores are plotted by mutational consequence. Data is shown for *n* = 3,843 SNVs assayed in regions outside the RING and BRCT domains. **C.** Function scores are plotted by genomic position for each new region assayed. Each SNV is coloured by mutational consequence. Black dashed lines represent the significance threshold (*q* = 0.01) for calling significantly depleted variants by region, while grey dashed lines (function score = −0.799) divide intermediate and LoF variants gene-wide.

We also performed SGE for two regions previously assayed, exons 5 and 17, and observed function scores to broadly correlate with published data (**Figs. S1B, S2**). Significantly depleted variants (*q* < 0.01) were defined for each SGE region. Depleted variants scoring comparably to nonsense variants gene-wide were deemed ‘loss-of-function’ (‘LoF’, function score < −0.799), whereas less strongly depleted variants were deemed ‘intermediate’. Variants not significantly depleted were deemed ‘neutral’. In newly assayed regions, 6.5% of variants tested scored as LoF and 1.9% of variants scored as intermediate, comprising a smaller fraction of functionally impactful variants compared to regions encoding the RING and BRCT domains (**Fig. S1C,D**).

### Mapping variant effects across regions of *BRCA1*

Variant effect maps (**Fig. 1C**) reveal consistent depletion of nonsense and canonical splice site variants with few exceptions. Notably, all variants at the exon 1 splice donor site scored neutrally. Exon 1 is non-coding, and this result is consistent with variants at the exon 2 acceptor site also scoring neutrally^7^.

All variants at the exon 5 donor site scored neutrally, in contrast to prior SGE results which showed these variants to be strongly depleted. This is important because variants in this region were previously assayed in the context of a synonymous variant, c.297G>A, included in experiments to block re-cutting by Cas9 (**Fig. S2A**). While c.297G>A is not predicted to impact canonical splicing^24^, it may prevent alternative splicing of a reported in-frame transcript missing nine nucleotides^25^. Therefore, it has remained unclear whether variants disrupting the canonical donor site lead to LoF when introduced without c.297G>A, as evidenced by conflicting interpretations in ClinVar. Here, we addressed this by performing SGE with a re-designed exon 5 HDR library lacking c.297G>A. This revealed no variants at the donor site or further into intron 5 to be LoF (donor site function scores: −0.30 to 0.14; **Figs. 1C, S2B–D**). This indicates that the previous SGE data for exon 5 were confounded by the use of c.297G>A to block re-cutting, and that the new data should be given precedence for variant interpretation.

### Missense variants in the coiled-coil domain are strongly depleted in HAP1

Apart from exons 5 and 17, which encode parts of the RING and BRCT domains, respectively, LoF missense variants are primarily observed in regions of exons 11 and 12 encoding BRCA1’s CC domain (**Fig. 2A**). Three variants within the CC domain were previously shown to disrupt the BRCA1–PALB2 interaction and lower BRCA1 HR activity^26^, with p.M1400V causing a partial HR reduction of 25%. Function scores for these three variants track closely with published HR activities.

Mapping function scores to the structure of the BRCA1 CC domain in complex with the PALB2 CC domain reveals interacting residues highly intolerant to missense variation (**Fig. 2B**). Missense variants at the *a* and *d* positions of BRCA1’s CC heptad repeat^27^ have a mean function score of −1.13, compared to −0.37 at other positions (Wilcoxon Rank-Sum *P =* 1.8×10^−5^). Likewise, CC-domain variants predicted by FoldX to destabilise the BRCA1– PALB2 interaction tended to have lower function scores (**Fig. 2C**; Spearman’s *ρ* = −0.54), indicating disruption of this interaction leads to LoF in HAP1.

### LoF variants outside structured domains impact splicing

To determine the extent to which SNVs outside BRCA1’s established structural domains cause LoF, we performed SGE of four regions of exon 10, as well as exon 6, in which variants have been reported to impact replication fork protection but not HR^23^. In exon 10, we prioritised two internal regions predicted by AlphaFold2^28^ to potentially encode alpha helices (**Fig. S3A**), as well as regions covering the 5’ and 3’ ends of the exon.

Apart from nonsense variants and variants in close proximity to splice sites, only four variants caused LoF across these regions: one synonymous and three missense variants. All four of these variants create new 5’-AGGT sequences near an established alternative splice donor site in exon 10^29,30^, have low RNA scores (range: −4.44 to −2.59) and are predicted by SpliceAI^24^ to create splice donor sites (**Fig. 2D,E**), indicating splice disruption to be the mechanism leading to LoF. Importantly, not a single variant was deemed LoF among 1,043 exon 10 missense variants that had RNA scores above −2.0. Likewise, all missense variants in exon 6 scored neutrally, including two variants reported to impact replication fork protection^23^.

### Non-coding variants impact function via multiple mechanisms

Thus far, few pathogenic *BRCA1* variants have been found in non-coding regions, apart from those adjacent to splice junctions^31,32^. To explore the potential of non-coding variants to cause LoF, we performed SGE of the proximal promoter and 5’-UTR using two overlapping HDR libraries (**Fig. 1A**). Low function scores were observed at highly conserved nucleotides (**Fig. S3B**) and highlighted a previously described 8-bp E2F transcription factor binding site^33^ (**Fig. 2G**). The functional importance of this binding site was confirmed by depletion of overlapping 1-and 5-bp deletions (**Fig. 2H**). Within the site, SNVs’ predicted effects on transcription factor binding correlated well with function scores (Pearson’s *r* = 0.57, **Fig. 2I**), corroborating the importance of this binding site in HAP1.

**Figure 2:**
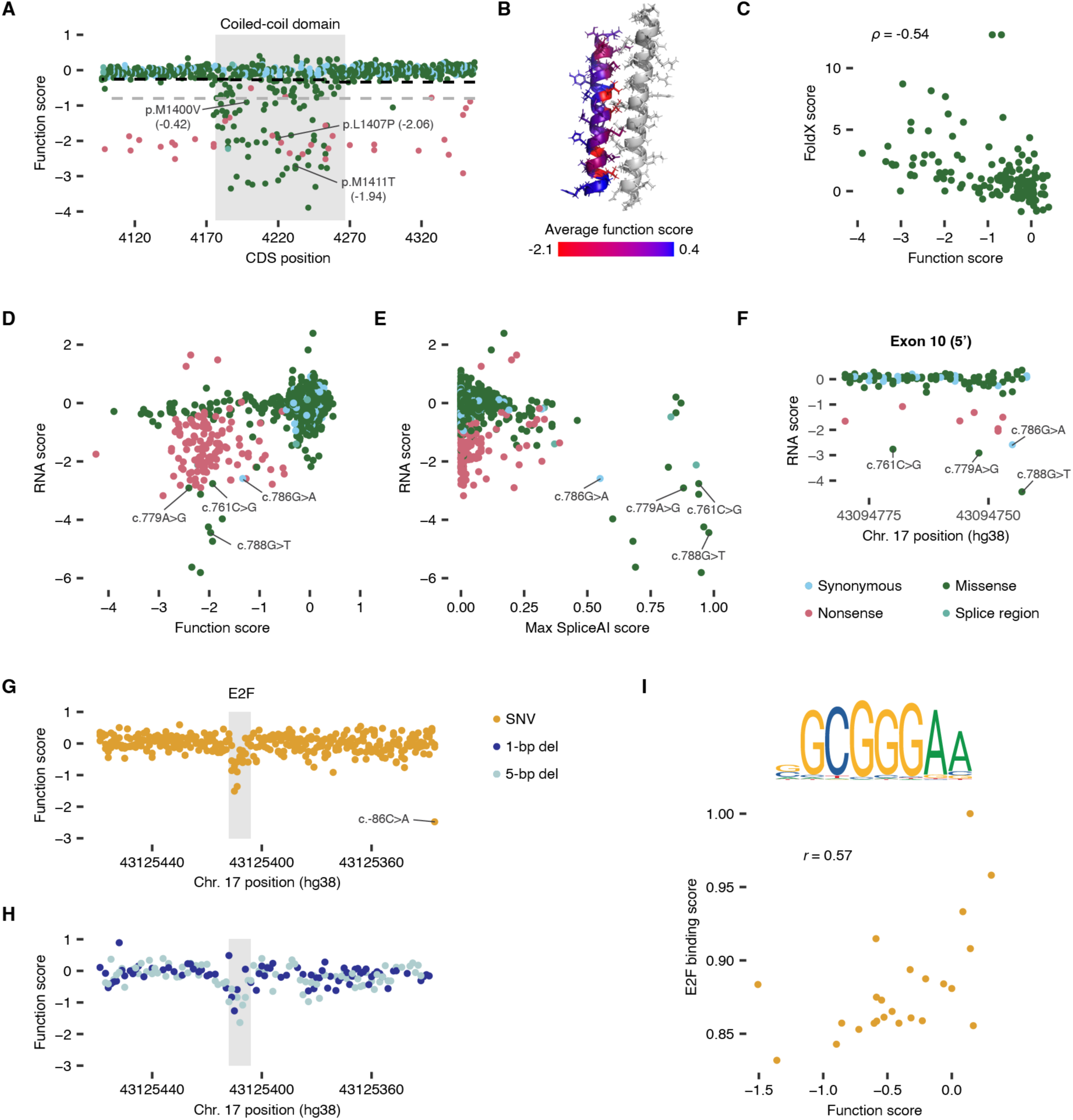
Newly identified LoF variants impact function by diverse mechanisms. **A.** Function scores are shown for *n* = 770 coding SNVs in exons 11 and 12, coloured by consequence. LoF missense variants occur almost exclusively within the CC domain, which is encoded by the region shaded in grey. Black and grey dashed lines show thresholds dividing neutral and intermediate, and intermediate and LoF categories, respectively. Missense variants shown by Anantha et al. to reduce HR activity^26^ are labelled, with the reported log_2_-fold change in HR activity shown. **B.** The mean function score for missense SNVs at each amino acid position is mapped onto the mouse protein structure of the BRCA1 CC domain in complex with PALB2’s CC domain in grey (PDB: 7K3S). **C.** Function scores are plotted by FoldX ΔΔG scores for *n* = 163 SNVs in the CC domain (Spearman’s *ρ* = −0.54). **D.** Function scores are plotted against RNA scores for each exonic SNV (*n* = 2,472). Four exon 10 variants predicted to impact splicing are labelled. **E.** RNA scores are plotted against maximum SpliceAI scores for the same exonic variants as in **D**. **F.** RNA scores are plotted by genomic position for *n* = 113 SNVs near the 5’ end of exon 10. One synonymous and three missense variants with low function scores and RNA scores are labelled. **G,H.** Function scores for *n* = 365 SNVs (**G**) and *n* = 270 1- and 5-bp deletions (**H**) in part of the promoter and 5’-UTR are plotted by genomic position. Low function scores coincide with an E2F binding site^33^, highlighted in grey. **I.** In the promoter and 5’-UTR regions, JASPAR predicts a binding site for DP1, which binds DNA in complex with E2F proteins. The 8-bp consensus binding motif MA1122.1 is shown. JASPAR-predicted binding scores for DP1 are correlated with function scores for SNVs within the 8-bp consensus sequence.

In the 5’-UTR, a single strongly depleted variant, c.−86C>A, was present in two overlapping libraries and scored as LoF in both (function scores = −2.48, −2.87). This variant results in a 5’-ATG sequence, thus creating an open reading frame (ORF) that overlaps the canonical ORF (**Fig. 2G**).

To explore the potential of SGE to identify pathogenic variants deep within introns, we targeted a region of intron 11 containing c.4185+4105C>T, a likely pathogenic variant in ClinVar. This variant is reported to create a splice donor site, promoting inclusion of a pseudoexon that leads to nonsense-mediated decay^32^. Among 388 SNVs tested in this region, c.4185+4105C>T was 1 of 2 SNVs to score as LoF (function score = −1.37; **Fig. S3C**), corroborating its pathogenicity. A nearby variant, c.4185+4108C>G, scored lower (−2.32). While c.4185+4108C>G has not been reported, like c.4185+4105C>T, it is predicted by SpliceAI to potentially disrupt splicing (maximum SpliceAI score = 0.29; **Fig. S3D**). This experiment, therefore, establishes that SGE can successfully discriminate pathogenic variants deep within introns, while also highlighting the role of computational predictors for prioritising such variants for functional study.

### Development of an SGE assay in mammary epithelial cells

To explore whether *BRCA1* variant effects are consistent across cell models, we developed an SGE assay in HMECs. HMECs are diploid, non-transformed cells of human mammary origin that may be a better model for breast cancer’s cell of origin. To enable delivery of a single variant per cell, we first knocked out one complete *BRCA1* allele by engineering an 81-kb deletion in an HMEC line with inducible Cas9 expression^34^ (**Fig. S4A**). Frameshifting insertions and deletions (indels) introduced to the remaining *BRCA1* allele were depleted over time, revealing an essential role of BRCA1 in HMEC growth (**Fig. S4B**).

To perform SGE of 10 *BRCA1* regions, we co-transfected HMECs with gRNA plasmids and corresponding HDR libraries after inducing Cas9 expression with doxycycline (**Fig. 3A**). To increase HDR editing, transfected cells were treated with NU7441, a DNA-PK inhibitor (**Fig. S4C**). Edited cells were harvested for amplicon sequencing on days 7, 14 and 21. Separate function scores were calculated using data from untreated HMECs and HMECs maintained in olaparib from day 7. Smaller libraries were used for technically challenging regions with few LoF variants in HAP1 (i.e., the promoter, intron 11, and exon 6) to ensure accurate scoring. In total, 2,157 variants were scored in HMECs (**Fig. 3B**).

**Figure 3.**
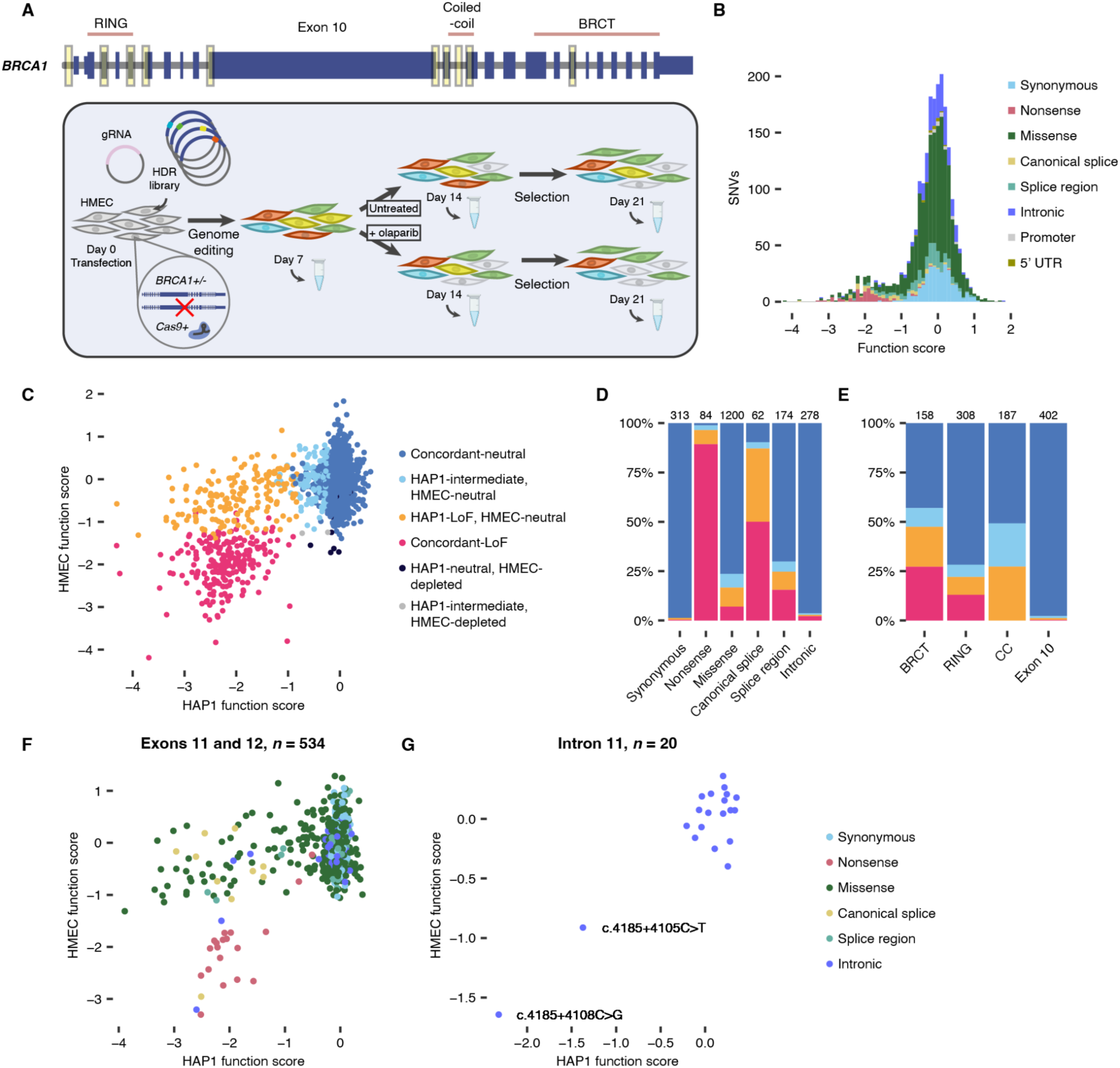
Comparing variant effects across cell lines using SGE in human mammary epithelial cells. **A.** Ten *BRCA1* regions highlighted in yellow were assayed in the HMEC line. The schematic shows the SGE protocol in *BRCA1*^+/−^ HMECs. After induction of Cas9 expression, cells are transfected with a gRNA-expressing plasmid and HDR library, then sampled and sequenced on days 7, 14 and 21. Function scores are calculated for both untreated and olaparib-treated cells. Experiments were performed in duplicate. **B.** The distribution of function scores from untreated HMECs is shown for variants across all ten *BRCA1* regions, coloured by mutational consequence (*n* = 2,157). **C.** HAP1 function scores are plotted by untreated HMEC function scores for the same SNVs as in **B**. SNVs are coloured by combined function class. For comparison, HAP1 scores for exon 3 were taken from Findlay et al.^7^ and normalised such that the median synonymous and nonsense scores match those in the new HAP1 dataset. **D,E.** The proportion of variants in each combined class is shown for each mutational consequence (**D**) and for missense variants in each BRCA1 protein domain (**E**). 11 variants depleted in HMEC and neutral or intermediate in HAP1 are not shown. **F,G.** HAP1 function scores are plotted against untreated HMEC function scores for all variants in the SGE regions of exons 11 and 12, which encode the CC domain (**F**), and intron 11 (**G**).

HMEC-derived function scores were well correlated across replicate transfections (**Fig. S4D**) and nonsense variants were strongly depleted compared to synonymous variants (mean function score −1.91 versus 0.02, respectively; **Fig. S4E**). Similarly to before, we used a false discovery rate (FDR) of 0.01 to identify 235 of 2,157 variants (10.9%) that were significantly depleted (**Figs. S4F, S5**).

### Comparing variant effects across cell lines

We next compared HAP1 function scores with HMEC function scores (**Fig. 3C**). While most variants were neutral in both assays, of 393 variants deemed LoF in HAP1, only 224 (57.0%) were significantly depleted in HMEC. Furthermore, only two of 100 (2.0%) HAP1-intermediate variants were depleted in HMEC. Meanwhile, only nine variants were depleted in HMEC but neutral in HAP1. We defined combined function classes for variants assayed in both lines, classifying 99.5% of variants as either ‘concordant-neutral’, ‘concordant-LoF’, ‘HAP1-LoF, HMEC-neutral’, or ‘HAP1-intermediate, HMEC-neutral’.

Many missense variants that scored as LoF or intermediate in HAP1 scored as neutral in HMECs (**Fig. 3D**). Discordantly classified missense variants were observed across protein domains (**Fig. 3E**). Notably, in the CC domain, all HAP1-LoF missense variants scored neutrally in HMECs (**Fig. 3F**). A large proportion of HAP1-LoF splice variants were also neutral in HMECs (**Fig. 3D**). For example, canonical splice variants at the exon 12 acceptor site scored neutrally in HMEC, suggesting that an alternative acceptor site leading to an in-frame transcript^35^ is able to preserve BRCA1 function in HMEC (**Fig. 1C**, **Fig. S5**). Exon 10 can be alternatively spliced, resulting in two in-frame isoforms (Δ10 and Δ10q)^29,35^. Exon 10 canonical splice site variants were significantly depleted only in HAP1 (**Fig. 1C, Fig. S5**). Likewise, among variants near the Δ10q splice junction that were LoF in HAP1, those predicted by SpliceAI to create in-frame transcripts scored neutrally in HMECs. Meanwhile, the HAP1-LoF variants deep within intron 11 scored highly similarly across lines (**Fig. 3G**). In contrast, the few HAP1-LoF variants identified in the 5’-UTR and promoter scored as neutral in HMECs.

**Figure 4.**
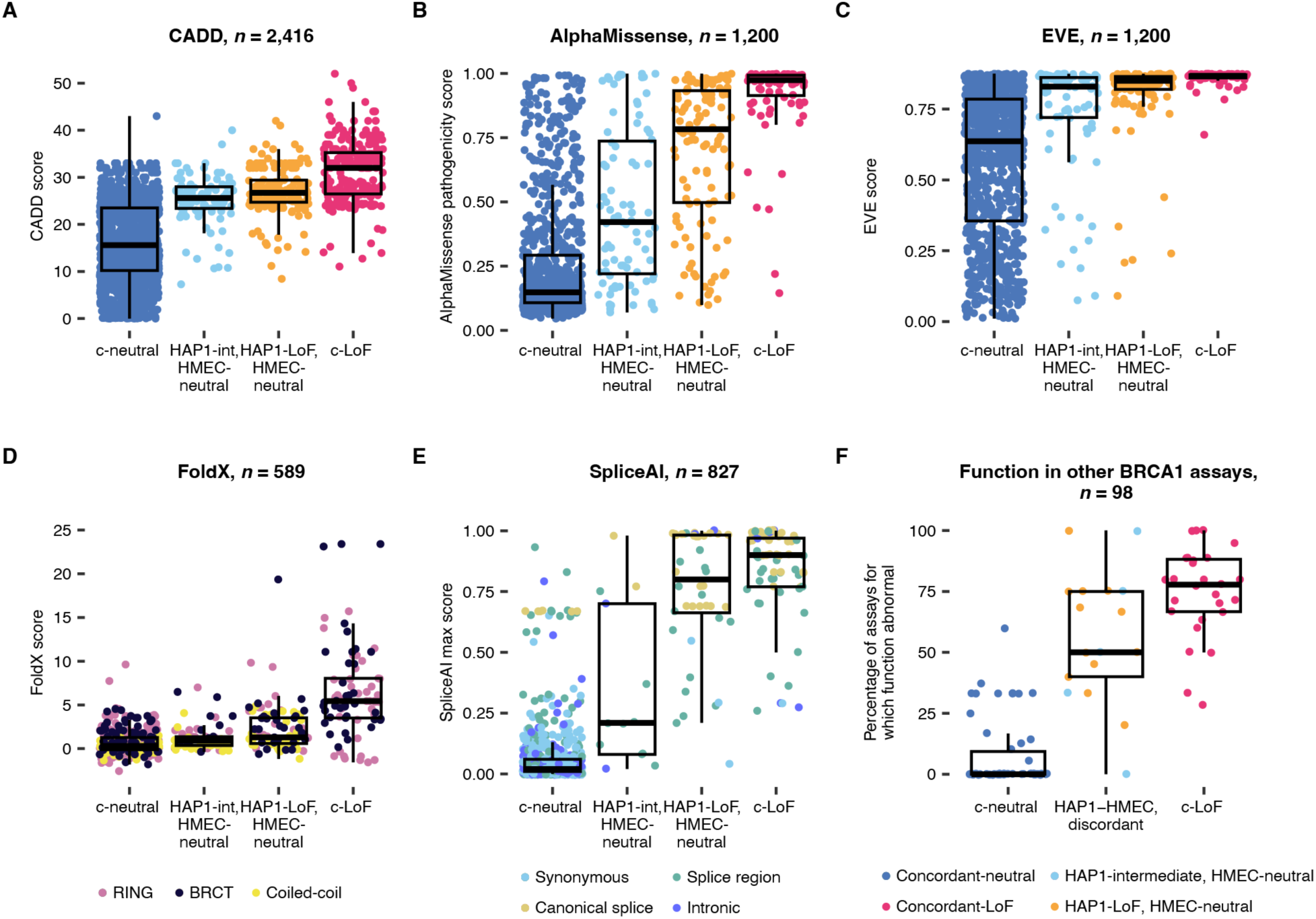
Discordantly scored variants are less deleterious to *BRCA1* function than concordant-LoF variants. **A–C.** For variants in each combined function class, scores from CADD (**A**, all SNVs), AlphaMissense (**B**, missense SNVs), and EVE (**C**, missense SNVs) are plotted. **D.** FoldX-predicted ΔΔG values are plotted for missense variants in the RING domain (*n* = 262), BRCT domain (*n* = 145) and CC domain in complex with PALB2 (*n* = 182). Variants predicted to impact splicing (maximum SpliceAI score > 0.2) were excluded. **E.** The maximum SpliceAI score for each variant is plotted by combined function class for intronic and synonymous variants. **F.** A total of 98 variants assayed in both HAP1 and HMEC had published data available from at least 3 assays measuring BRCA1 function^18^. For each variant, the percentage of assays that deemed the variant to be functionally abnormal is plotted by combined function class. ‘HAP1–HMEC discordant’ includes variants classed as LoF or intermediate in HAP1, and as neutral in HMECs. (Prior SGE data from Findlay et al. was excluded from this analysis.) (Box plots: centre line, median; box limits, upper and lower quartiles; whiskers, 1.5× interquartile range; all points shown.)

Across variant effect predictors including CADD, AlphaMissense, EVE, FoldX and SpliceAI, concordant-LoF variants were predicted to have the strongest effects (**Fig. 4A–E**). For instance, whereas concordant-LoF missense variants had a median AlphaMissense score of 0.97, the median score for HAP1-LoF, HMEC-neutral variants was 0.78 (Wilcoxon Rank-Sum *P =* 1.2×10^−13^). Likewise, concordant-LoF missense variants in structured domains were predicted by FoldX to be much more destabilising (median concordant-LoF ΔΔG = 5.44 versus median HAP1-LoF, HMEC-neutral ΔΔG = 1.29; Wilcoxon Rank-Sum *P* = 2.5×10^−11^). Analysing published data^18^, we also found discordantly classified variants to be less consistently scored in other assays (**Fig. 4F**). These results confirm concordant-LoF variants to be more highly deleterious than variants scoring as LoF or intermediate in HAP1 but neutral in HMECs.

### Functional redundancy underlies differential effects across lines

We next explored potential mechanisms underlying differences between HAP1 and HMEC. *BRCA1* variants disrupting HR sensitise cells to PARP-inhibitor treatment^36^. We therefore analysed 1,464 variants in HMECs treated with olaparib to ask whether additional variants were depleted (schematic: **Fig. 3A**). Function scores from untreated and olaparib-treated HMECs were highly correlated overall (**Fig. 5A**). However, 61 HMEC-neutral variants selectively scored as LoF upon addition of olaparib. Among these, 78.7% also scored as LoF in HAP1. Variants significantly depleted in HMEC only upon PARP inhibition include variants at the exon 10 splice donor site (**Fig. 5B,C**) and 25 CC-domain missense variants (**Fig. S6A**).

Given the importance of the BRCA1–PALB2 interaction in facilitating RAD51 loading during HR, we examined PALB2’s essentiality in HMEC. Frameshifting indels were strongly depleted when *PALB2* was targeted using CRISPR (**Fig. S6B**), establishing the gene’s essentiality. One mechanism by which PALB2 can be recruited independently of BRCA1 during HR is via direct association with the E3 ligase RNF168^37^. Unlike PALB2, RNF168 was not essential in HMECs with intact BRCA1 function (**Fig. S6C**).

To investigate the possibility that RNF168 may functionally compensate for variants disrupting the BRCA1–PALB2 interaction in HMECs (**Fig. 5D**), we created an RNF168^−/−^ HMEC line and again performed SGE to assay CC-domain variants in exon 12. In RNF168^−/−^ HMECs, many CC-domain missense variants scored more lowly than in parental HMECs, both with and without olaparib (**Fig. 5E,F**). Indeed, many missense variants deemed LoF in HAP1 but neutral in parental HMECs scored as lowly as nonsense variants in the RNF168^−/−^ line. These experiments indicate that RNF168 provides at least partial compensation for disruption of the BRCA1–PALB2 interaction in HMECs.

**Figure 5.**
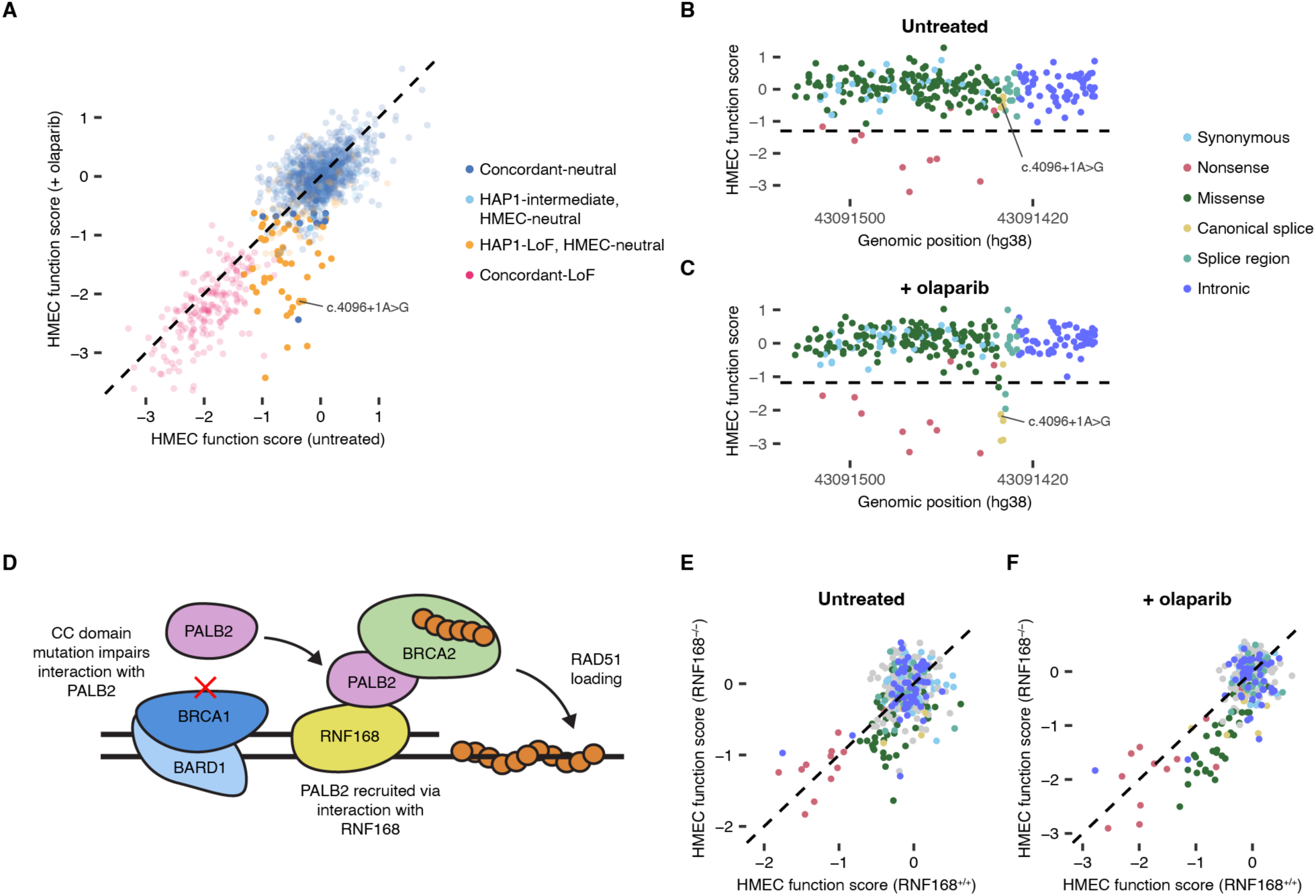
Mechanisms underlying differential variant effects across cell lines. **A.** HMEC function scores are shown for untreated compared to olaparib-treated cells for *n* = 1,438 variants, coloured by combined function class as defined using untreated scores. Variants represented by opaque points scored as depleted in HMEC only when treated with olaparib. **B,C.** Function scores for SNVs in the 3’ exon 10 region are shown by position for untreated (**B**, *n* = 261) and olaparib-treated (**C**, *n* = 262) HMECs. **D.** Schematic showing a coil-coiled domain missense variant impairing the BRCA1–PALB2 interaction. PALB2 can be alternatively recruited via RNF168, facilitating RAD51 loading onto resected DNA. **E,F.** SGE function scores for CC-domain variants in the parental HMEC line compared to the RNF168^−/−^ HMEC line, for untreated (**E**, *n* = 344) and olaparib-treated (**F**, *n* = 343) cells. Variants are coloured as in **B**, except for missense variants that were not LoF in HAP1, which are grey. Function scores are normalised such that the median nonsense and synonymous scores across cell lines match.

### Accurate discrimination of pathogenic variants

Across studies, SGE performed in a single cell line has successfully discriminated between known pathogenic and benign variants^7,9,10,12–14^. Our data present a unique opportunity, however, to ask whether SGE data from multiple lines can improve prediction of variant pathogenicity and cancer risk.

We first compared how well our individual HAP1 and HMEC datasets perform at classifying variants in accordance with ClinVar annotations. For this, we used the set of *n* = 160 pathogenic or likely pathogenic (P/LP) variants and *n* = 260 benign or likely benign (B/LB) variants with a minimum review status of one star that were assayed in both lines. Overall, the HAP1 assay performed better (HAP1 area under the receiver operating characteristic curve (AUC) = 0.997; HMEC AUC = 0.977; **Fig. 6A,B**). Performing the same analysis for all 581 one-star P/LP or B/LB variants assayed in HAP1 yielded a similarly high AUC of 0.997 (**Fig. S7A**).

The difference across lines stems largely from P/LP variants scoring as LoF or intermediate in HAP1 and as neutral in HMEC (**Fig. 6C**). Whereas 98.1% of all B/LB variants were concordant-neutral, only 81.3% of P/LP variants were concordant-LoF. Among P/LP variants not deemed concordant-LoF, 22 of 30 were HAP1-LoF, HMEC-neutral and 6 of 30 were HAP1-intermediate, HMEC-neutral. Most VUS were classified as concordant-neutral (72.8%). However, VUS were enriched among discordantly classified variants, comprising 34.9% of HAP1-LoF, HMEC-neutral variants and 30.6% of HAP1-intermediate, HMEC-neutral variants (**Fig. 6D**).

Functionally neutral *BRCA1* variants are more likely to be observed in population controls. Consistent with this, concordant-neutral variants were most frequent in the gnomAD^38^ and All of Us databases (**Fig. 6E,F**), followed by discordantly scored variants, which were more frequent than concordant-LoF variants.

### Predicting clinical risk from SGE data

We next examined whether discordantly scored variants may confer intermediate cancer risk. Previous SGE^7^ of exon 17 failed to score a *BRCA1* variant well-established to confer intermediate risk, p.R1699Q^39,40^. Here, we performed SGE of exon 17 using an HDR template encoding p.R1699Q via dinucleotide substitution (c.5096_5097delinsAA) to enable accurate scoring. Whereas p.R1699Q scored as lowly as nonsense variants in HAP1 (function score = −2.38), the same variant was not significantly depleted in HMEC (function score = −0.37; **Fig. S7B,C**).

A recent analysis indicated that SNVs impacting the exon 10 splice acceptor site may be pathogenic with reduced penetrance^41^. In HAP1, 5 of 6 such variants scored as LoF, while c.671-1G>A scored as intermediate (mean function score = −2.03). In contrast, 0 of 6 variants at this splice site were depleted in HMEC (mean function score = −0.85; **Fig. 6C**). Evidence from the ENIGMA consortium indicates that the exon 10 splice donor variant c.4096+1G>A may also be pathogenic but confer reduced risk^42,43^. Similarly to exon 10 acceptor site variants, c.4096+1G>A scored as HAP1-LoF, HMEC-neutral (HAP1 function score = −2.28, HMEC function score = −0.37; **Fig. 6C**). Notably, this variant was strongly depleted in HMECs treated with olaparib (function score −2.14; **Fig. 5A–C**). In summary, all eight variants reported to potentially confer intermediate cancer risk were significantly depleted in HAP1 but not untreated HMECs.

**Figure 6:**
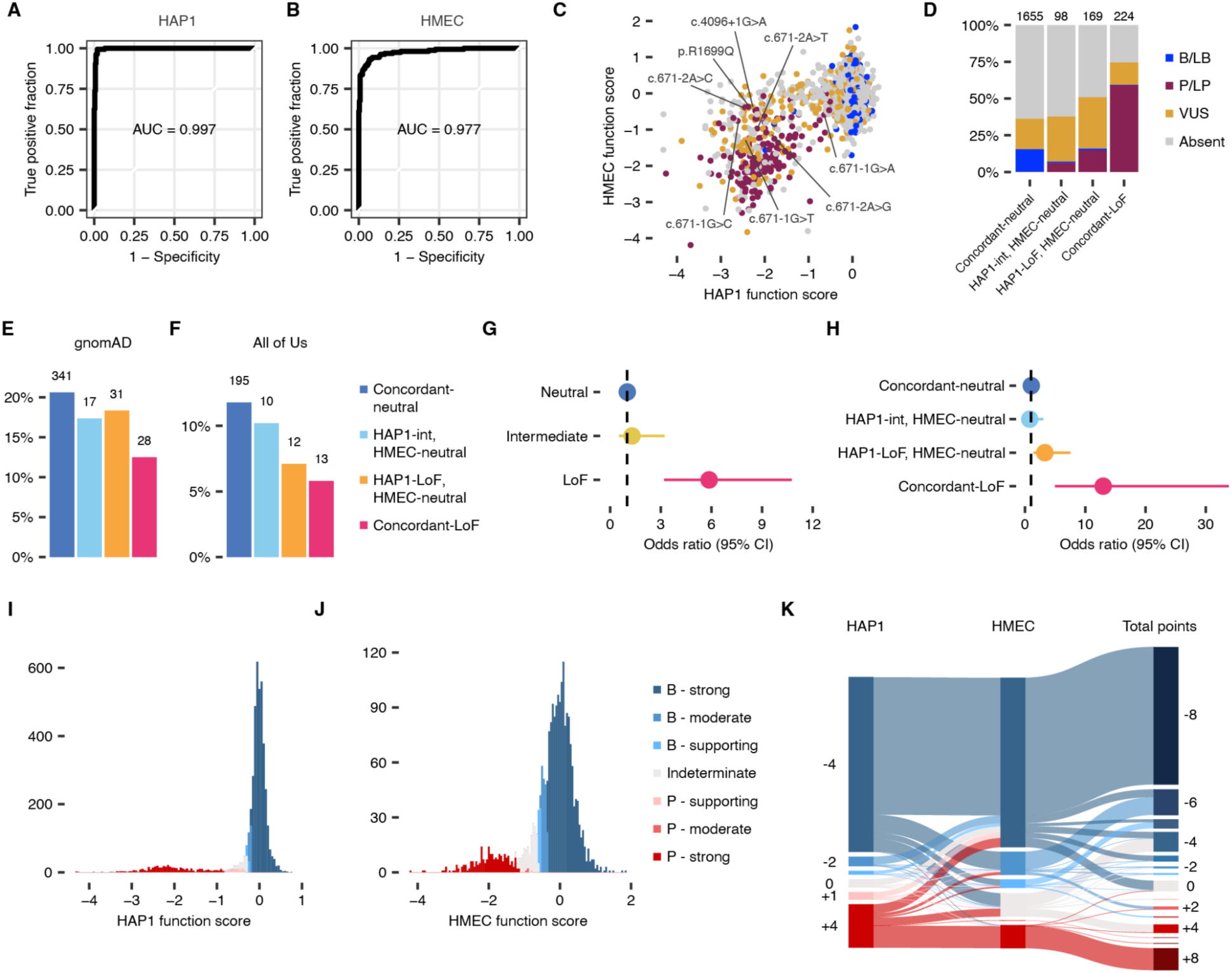
Refining clinical risk estimates of functionally abnormal variants. **A,B**. Receiver operating characteristic (ROC) curves compare the performance of HAP1 (**A**) and HMEC (**B**) function scores for classifying ClinVar B/LB (*n* = 260) and P/LP (*n* = 160) variants with a minimum review status of one star. **C.** HAP1 function scores plotted against HMEC function scores are shown for *n* = 2,157 variants, coloured by ClinVar interpretation. Labelled variants have been reported to be pathogenic with reduced penetrance. **D.** For each combined function class, the proportion of each ClinVar classification is shown. **E,F.** For each combined function class, the percentage of variants present in the gnomAD (**E**) and All of Us (**F**) databases are shown. **G,H.** Odds ratios (ORs) for breast cancer risk determined from meta-analysis of BRIDGES and UK Biobank case–control data^44^ are plotted for function classes defined using HAP1 data alone (**G**) or by combined data (**H**). Lines indicate 95% confidence intervals. **I,J.** The ACMG/AMP framework^45^ was applied to assign PS3 and BS3 strengths of evidence for *n* = 4,470 variants assayed in HAP1 (**I**) and *n* = 2,157 variants assayed in HMEC (**J**). The distribution of function scores for each assay is shown, coloured by the final evidence code assigned. **K.** Sankey plot showing evidence codes for *n* = 1,867 variants in the HAP1 and HMEC datasets. Total points were calculated by adding points from individual lines. (Data from exon 3 were excluded as HAP1 function scores were derived in previous SGE experiments^7^.)

Next, we estimated breast cancer risk for each function class using case–control data from 66,859 female breast cancer cases and 271,189 controls from the BRIDGES study of the Breast Cancer Association Consortium (BCAC)^44^ and the UK Biobank^46^. Odds ratios (ORs) with 95% confidence intervals (CIs) were estimated for each dataset and combined using a fixed-effects, inverse variance meta-analysis to derive overall association^47^, yielding similar findings across cohorts and in the combined dataset (**Table S5**). Of the three HAP1-based function classes, only variants deemed LoF were significantly associated with higher breast cancer risk (meta-analysis OR = 5.86, 95% CI = 3.19–10.75; **Fig. 6G**). This result remained significant when LoF variants in the RING and BRCT domain were excluded (OR = 4.35, 95% CI = 2.27–8.32; **Table S5**). Notably, HAP1-intermediate variants were not associated with significantly increased risk (OR = 1.29, 95% CI = 0.52–3.23).

We reasoned that function classes derived by combining HAP1 and HMEC data may provide improved resolution. Indeed, while HAP1-LoF, HMEC-neutral variants were associated with moderately elevated risk (OR = 3.28, 95% CI = 1.43–7.52), concordant-LoF variants were associated with higher risk (OR = 12.97, 95% CI = 4.97–33.82; **Fig. 6H, Table S5**). In contrast, both concordant-neutral variants and HAP1-intermediate, HMEC-neutral variants were not associated with increased risk. For the concordant-LoF class, a similarly high OR was obtained even when nonsense and canonical splice site variants were excluded from analysis (OR = 10.84, 95% CI = 2.50–46.99).

### Evidence for clinical variant interpretation

Lastly, we explored how the HAP1 and HMEC SGE datasets may best be leveraged to support clinical variant interpretation. First, for each dataset, we defined truth sets of pathogenic and benign variants using ClinVar annotations with at least two stars and fit a Gaussian mixture model (**Fig. S7D,E**) to determine the odds of pathogenicity (OddsPath)^48^ for each variant, as established^14^. This facilitated assigning evidence codes^45^ reflective of the strength of functional evidence from each cell line (**Fig. S7F,G**).

The majority of variants were assigned evidence codes consistent with their function class in both HAP1 and HMEC (**Tables S1, S3**). Notably, though, many HAP1-intermediate variants were initially assigned strong evidence in favour of pathogenicity. This is at odds with the fact that HAP1-intermediate variants were not associated with significantly increased breast cancer risk (**Fig. 6G**). A small proportion of variants not significantly depleted were also assigned evidence of pathogenicity in at least one cell line. Based on this, we adjusted evidence codes in two ways: 1.) evidence codes in favour of pathogenicity were replaced with ‘indeterminate’ for variants not significantly depleted (*q* > 0.01); 2.) strength of evidence in favour of pathogenicity was capped at ‘supporting’ for HAP1-intermediate variants. These adjustments impacted only 4.0% (268 of 6,627) of final evidence codes (**Fig. 6I,J**).

To examine the combination of functional evidence from the two SGE assays, we summed the points associated with individual evidence codes to obtain a combined points total for variants assayed in both lines (**Fig. 6K**). This revealed 12.3% (*n* = 230) of variants to receive at least 4 points in favour of pathogenicity and 77.8% (*n* = 1,452) of variants to receive at least 4 points in favour of benignity. Only 5.2% (*n* = 97) of variants did not receive at least 2 points in either direction. As guidelines for the integration of multiple lines of functional evidence continue to evolve, we aim to support future *BRCA1* variant interpretation by supplying all function scores and evidence codes in **Tables S1 and S3**.

## DISCUSSION

Here, we applied SGE to systematically characterise the functional effects of *BRCA1* variants across two cell lines, HAP1 and HMEC. We identified variants leading to LoF by diverse mechanisms, both in coding and non-coding regions, providing valuable evidence for clinical variant interpretation. The unique perspective afforded by assaying variants in cells of mammary origin in addition to HAP1 permitted a more precise accounting of *BRCA1*-related cancer risk.

### Identifying new LoF variants in *BRCA1*

Nearly all newly identified LoF missense variants in HAP1 map to the CC domain, implicating the BRCA1–PALB2 interaction as being crucial for HAP1 cell viability. While variants in the CC domain have previously been shown to impact function^26,49,50^, this is the first high-quality functional data for all possible CC-domain SNVs. Critically, in contrast to HAP1, no CC-domain missense variants were significantly depleted in untreated HMECs. This difference between lines was partially reconciled by olaparib treatment or knock-out of RNF168, suggesting the same set of missense variants in the CC domain impact BRCA1 function in both lines, but that effects on HMEC growth are, at most, subtle under normal conditions. One implication is that all missense variants disrupting the BRCA1–PALB2 interaction preserve other functions of BRCA1, including promotion of DNA end resection^51^. Further, these data suggest that pathogenic CC-domain missense variants are unlikely to cause the same degree of cancer risk as protein-truncating variants.

Only 8 of 1,665 missense variants assayed outside the RING, BRCT and CC domains were LoF in HAP1. This finding is consistent with an estimate from case–control data that only 1% of missense variants outside BRCA1’s structured domains are pathogenic^52^. All 8 LoF variants identified had SpliceAI scores above 0.60, suggesting impairment at the level of pre-mRNA splicing rather than protein function, a finding confirmed by low RNA scores in SGE (range: −5.62 to −2.19). Among neutrally scored missense variants were 1,037 SNVs in exon 10. Exon 10 comprises ∼60% of *BRCA1* coding sequence and accounts for 2,522 missense VUS in ClinVar, most of which remain unassayed. Our work provides strong experimental support for considering exon 10 missense variants without predicted effects on splicing to be likely benign rather than VUS, as has been proposed^53^.

Beyond coding regions, both the HAP1 and HMEC assays correctly identified a pathogenic deep intronic variant known to disrupt splicing, c.4185+4105C>T, demonstrating the ability of SGE to identify pathogenic variants in deep intronic regions (**Fig. 3G**). It is notable that c.4185+4105C>T and an unreported LoF variant nearby, c.4185+4108C>G, both had SpliceAI scores greater than 0.20 (0.27 and 0.29, respectively), whereas other variants in the region did not. Additional pathogenic deep intronic variants in *BRCA1* have recently been reported^54^ and case–control analysis has revealed a significant excess of deep intronic germline variants in breast cancer cases^52^. Going forward, therefore, we expect computational prioritisation of variants impacting splicing together with functional testing will prove highly valuable for identifying additional pathogenic variants in *BRCA1*.

HAP1-depleted variants in the promoter and 5’-UTR predicted to disrupt transcription factor binding and translation initiation scored neutrally in HMECs. While a previous study linked c.−107A>T to promoter hypermethylation and reduced *BRCA1* expression^31^, this variant did not score significantly in either line. Therefore, caution is warranted in using SGE data derived in HAP1 to adjudicate promoter and 5’-UTR variants, as their effects may be cell-type specific.

### Leveraging data across cell lines to improve variant classification

By developing an SGE protocol in a non-transformed line of mammary origin, we were able to systematically compare its classification performance to that of HAP1-based SGE. Both lines distinguished pathogenic variants with high specificity, but HAP1 showed better overall performance despite being the product of an unsuccessful attempt to induce pluripotency in cells of myeloid origin^55^. Importantly, though, rather than confounding variant interpretation, our analyses revealed clear benefits to scoring variants in multiple lines.

Firstly, 79.3% of variants tested in both lines were assigned evidence codes concordantly in support of pathogenicity or benignity. For such variants, the HMEC function scores strengthen the functional evidence available for classification. Although we capped functional evidence from a single line at ‘strong’ (equivalent to four points), 78.9% of variants reached at least five points in either direction once evidence was summed.

Second, it has been challenging to identify pathogenic variants that confer reduced cancer risk compared to protein-truncating variants. For this task, the HMEC assay proves highly valuable. As a single class, HAP1-LoF variants were estimated to increase breast cancer risk 5.86-fold. However, HAP1-LoF variants also supported by a LoF classification in HMEC were associated with a 12.97-fold risk increase, whereas HAP1-LoF variants scored neutrally in HMEC were associated with only a 3.28-fold risk increase. This indicates that the two SGE assays can be used together to identify variants of reduced penetrance. Indeed, of eight variants tested that have been linked to reduced penetrance, including the well-characterised p.R1699Q, all eight were depleted in HAP1 but not HMEC.

Lastly, when initially assigning evidence strengths to function scores from HAP1, all intermediate variants with function scores below −0.42 initially received strong evidence in favour of pathogenicity (PS3). We found this to be unwarranted, as HAP1-intermediate variants as a class were not associated with significantly higher cancer risk. A recent HAP1-based SGE analysis of *RAD51C* similarly found intermediately scoring variants to not associate with increased cancer risk^10^. These results highlight the importance of calibrating the complete spectrum of functional effects to clinical risk. As functional assay calibration typically relies on firmly established pathogenic and benign variants, variants of intermediate effect are likely to be poorly represented. Here, only two of 100 HAP1-intermediate variants were depleted in HMEC, demonstrating how SGE data from multiple cell types can improve interpretation of intermediately scored variants.

Our study has several limitations. Despite optimisation, lower HDR rates in HMECs resulted in function scores lacking the same precision as those derived using the well-optimised HAP1 SGE assay. While the HMEC assay’s value is reflected by its success at distinguishing pathogenic and benign variants, the lower precision precluded defining variants of intermediate effect. Furthermore, while we demonstrated that PARP inhibition and knock-out of RNF168 in HMECs lead to more LoF variants scoring concordantly across lines, not all cell line-specific effects were fully reconciled. As discordantly scored variants were observed across BRCA1 domains, it is likely that multiple mechanisms underlie differences between lines, consistent with BRCA1 playing multiple roles in HR. Future studies investigating how discordantly scored variants impact specific functions of BRCA1 will prove valuable for further disentangling mechanisms. Lastly, although our estimates of cancer risk by function class were consistent across independent cohorts, more granular risk estimation will require larger sample sizes.

In conclusion, this study provides new insights into *BRCA1* variants that impact function outside of the well-studied RING and BRCT domains, substantially expanding our ability to apply functional evidence to clinical *BRCA1* variant classification. By demonstrating that variant effects often differ substantially between cell lines, we highlight how multiple SGE assays can be leveraged to more precisely predict cancer risk.

## METHODS

### HDR library design and cloning

Each SGE experiment requires a plasmid expressing a gRNA that cuts within the region to be mutated, and an HDR plasmid library that provides a repair template for each SNV. Where possible, libraries were designed to be compatible with two independent gRNAs. gRNAs were designed using the CRISPR design tool in Benchling. For each SGE region, one or two PAM-blocking edits not impacting the amino acid sequence were included to prevent Cas9 recutting after successful HDR editing, and to distinguish true HDR editing from sequencing error during analysis. Where it was not possible to mutate a given PAM sequence with a synonymous substitution, one to two mutations within the gRNA seed region were used instead. A script was used to generate sequences with every possible SNV spanning the SGE region, as before^12^, and additional sequence was added to each end to allow PCR amplification. For 5’-UTR and promoter SGE regions, 1-bp and 5-bp deletions at every position were included in addition to SNVs.

Oligos were ordered in a pool from Twist Bioscience. Each SGE region was amplified by PCR and purified using AMPure XP (Beckman Coulter). For each SGE region, a homology arm (HA) plasmid was created by PCR-amplifying a genomic fragment including 500–1,000 bp of sequence on each side of the SGE region and cloning this fragment into a linearised pUC19 vector using In-Fusion cloning (Takara Bio). Each HA plasmid was then linearised by PCR, using primers to create 15–20 bp of overlap with the corresponding oligo pool amplification product. Linearised vectors were treated with DpnI (NEB) to remove circular plasmid followed by purification. In-Fusion cloning was used to clone the amplified oligo libraries into the linearised HA plasmids and the resulting products were used to transform Stellar Competent Cells (Takara Bio). To check efficiency of the transformation, 1% of each transformation was plated onto ampicillin-containing plates, with the rest cultured overnight in 150 ml luria broth (LB) containing 100 µg/ml carbenicillin. From these cultures, final HDR libraries were purified using the ZymoPure Maxiprep kit (Zymo Research).

The original HDR library for exon 17 from Findlay et al.^7^ did not contain a SNV encoding p.R1699Q (c.5096G>A) because it overlapped with a PAM-blocking edit included in library design. We therefore designed a 2-bp variant (c.5096_5097delinsAA), which encodes p.R1699Q and is predicted to block re-cutting. An HDR template encoding this variant was made using *in vivo* assembly^56^ to modify the exon 17 HA plasmid. This was then mixed with the original HDR library at a molar ratio of approximately 1:100 for use in SGE.

### Cloning gRNA-expressing plasmids

For experiments in HAP1 cells, gRNAs were cloned into pX459 by ligating annealed oligos into BbsI-digested pX459 plasmid^57^. For SGE experiments in HMECs, Cas9 was not required in the gRNA-expressing plasmid and puromycin selection could not be used as the puromycin resistance gene was introduced when the cell line was immortalised. Therefore, gRNAs were cloned into pGL3-U6-sgRNA-PGK-blasticidin, which was derived from pGL3-U6-sgRNA-PGK-puromycin^58^ (Addgene #51133) using In-Fusion cloning. Cloning of individual gRNAs proceeded using the same method as for pX459, except that pGL3-U6-sgRNA-PGK-blasticidin was digested with BsaI, and the 5’ overhangs used on gRNA primers were 5’-CCGG for the forward primer and 5’-AAAC for the reverse primer. All gRNA plasmids were verified by Sanger sequencing.

### HAP1 cell culture

HAP1 cells with a frameshifting *LIG4* mutation^7^ were cultured in IMDM (Gibco) completed with 10% FBS and 1% penicillin-streptomycin (Gibco). To avoid reversion to diploid, cells were cultured in 2.5 µM 10-deacetyl-baccatin-III (DAB; Stratech) and maintained below 80% confluency. To passage, cells were washed twice in Dulbecco’s Phosphate Buffered Saline (DPBS; Gibco), trypsinised with 0.25% trypsin-EDTA (Gibco), neutralised with complete IMDM, centrifuged at 300 rcf for 5 min and resuspended.

### HAP1 SGE protocol

The day before transfection, 18 million cells were seeded onto a 10-cm tissue culture plate. On the day of transfection (day 0), cells were washed twice in DPBS and media was replaced without DAB. Cells were transfected using Xfect (Takara Bio), according to the manufacturer’s protocol with the following modifications: 30 µg gRNA-expressing plasmid, 10 µg HDR library and 24 µl Xfect polymer were combined in a total volume of 800 µl per transfection of each 10-cm plate. A negative control using the HDR library and a pX459 plasmid expressing a gRNA targeting *HPRT1*, a non-essential gene in HAP1, was performed for each experiment. The same transfection conditions were used but scaled down proportionally to a 6-well plate. On day 1, cells were washed, trypsinised and plated in a 15-cm plate with 1 µg/ml puromycin (Cayman Chemical). On day 4, cells were passaged; puromycin was removed and DAB was added to the media. Cells were then passaged on days 5, 8 and 11, with at least 6 million cells being plated each time. On days 5 and 14, pellets of up to 10 million cells were collected and stored at −80 °C. Negative controls were harvested on day 5.

### HMEC cell culture

hTERT-immortalised human mammary epithelial cells (HMECs) with doxycycline-inducible Cas9 expression^34^ were cultured in HuMEC medium (Gibco) with 2% FBS and 1% penicillin-streptomycin. To passage, cells were washed in DPBS, trypsinised with 0.25% trypsin-EDTA (Gibco), neutralised with Defined Trypsin Inhibitor (Gibco), centrifuged at 180 rcf for 5 min and resuspended in media.

### Generation of a *BRCA1*^+/−^ HMEC line for SGE

As the HMEC line is diploid, to ensure only one variant would be edited into the genome of each cell in SGE experiments, we deleted one *BRCA1* allele by using two gRNAs to simultaneously cut in non-coding regions upstream and downstream of *BRCA1*. One day before transfection, cells were plated in 12-well plates with 1 µg/ml doxycycline (Sigma-Aldrich) to induce Cas9 expression. Cells were transfected with 25 nM of each crRNA:tracrRNA complex (Edit-R, Horizon Discovery) using DharmaFECT 4 (Horizon Discovery) according to the manufacturer’s protocol. Doxycycline was removed from the media on day 2 and cells were expanded before limiting dilution was used to generate clonal lines. To identify lines with a *BRCA1* deletion, gDNA was extracted by adding 30 µl QuickExtract (Biosearch Technologies) to each well of a 96-well plate, transferring contents to a PCR plate and heating to 65 °C for 6 min and 98 °C for 2 min in a thermocycler. PCR with 1 µl template was then performed to identify lines with a complete *BRCA1* deletion using primers spanning *BRCA1*, and to check for mutations on the non-deleted allele using primers spanning each cut site. PCR products were purified using AMPure XP beads and Sanger sequenced. A cell line with complete deletion of one *BRCA1* allele and a 1-bp deletion upstream (NC_000017.11:g.43125556del) and a 1-bp insertion downstream (NC_000017.11:g.43044294_43044295insG) on the remaining allele was selected for use in SGE experiments.

### HMEC saturation genome editing

The day before transfection, 4.2×10^5^ cells were seeded per well with 1 µg/ml doxycycline (Sigma-Aldrich). Transfection was performed in 12-well plates using 12–24 wells per replicate. On the day of transfection (day 0), cells were washed with DPBS and 500 µl media containing doxycycline was added per well. Cells were transfected using Xfect; 1.5 µg of HDR library, 1.5 µg gRNA-expressing plasmid and 1.2 µl Xfect polymer were used in a volume of 50 µl per well. Three wells were used for negative controls. After 6 hours, cells in wells from the same replicate were pooled and plated between two 15-cm plates. To increase HDR editing efficiency, the DNA-PK inhibitor NU7441 (Stratech) was added to either 300 nM (SGE regions: exons 6, 11 and 10 (3’), intron 11) or 600 nM (SGE regions: promoter, exons 3 and 17). Blasticidin (Gibco) was included in the media from days 1–5 at 8 µg/ml, with cells being trypsinised and plated onto a fresh 15-cm plate on day 4 to aid selection. On day 7, doxycycline and NU7441 were removed from the media. Half the cells were harvested and half were divided between two 15-cm plates: one plate was treated with 300 µM olaparib and the other left untreated. The addition of olaparib was included in all HMEC SGE experiments apart from exon 5 and the 5’ exon 10 region. For the 5’ exon 12 region, untreated and olaparib-treated experiments were performed using two separate transfections. Cells were split on days 11, 14, 18 and 21, with 3 million cells plated per 15-cm plate. Up to 10 million cells were harvested on days 14 and 21.

### Nucleic acid extraction

Cell pellets were thawed at room temperature and homogenised using QIAshredder columns (Qiagen). Genomic DNA and RNA were extracted using the AllPrep DNA/RNA Kit (Qiagen), according to the manufacturer’s instructions. Genomic DNA was eluted in 100 µl nuclease-free water and quantified using the Qubit BR DNA Kit (Thermo Fisher Scientific). For HAP1 SGE samples from day 5, RNA was also extracted and then quantified using Nanodrop UV spectrometry.

### PCR1: amplifying gDNA

Each SGE region was amplified from gDNA, using at least one primer outside of the homology arms to prevent amplification of any HDR library still present in the samples. Primers were tested and annealing temperatures optimised using gradient PCR performed on unedited DNA. Up to 2 µg gDNA was used per 100 µl PCR reaction, with up to 8 reactions per sample, apart from the negative control, for which only one reaction was performed. PCR was performed using KAPA HiFi HotStart ReadyMix (Roche), with MgCl_2_ added to a final concentration of 5 mM, on a CFX96 Deep Well Real-Time PCR (Bio-Rad). SYBR Green (Thermo Fisher Scientific) was added to all PCR reactions at 0.4× to allow real-time reaction monitoring and stopping upon completion. After amplification, replicate reactions for each sample were pooled and examined by gel electrophoresis.

### PCR2: adding Nextera adapters

A second PCR was used to add Nextera sequencing adapters onto the amplicons from PCR1. Each 25 µl PCR2 reaction used 1 µl of pooled PCR1 product as a template and was run for 8–10 cycles. PCR products were purified using AMPure XP.

### cDNA preparation and amplification

For HAP1 day 5 samples, cDNA was prepared from 5 µg RNA using SuperScript IV First-Strand Synthesis (Invitrogen) using a *BRCA1*-specific primer. cDNA was then amplified as described for PCR2 using primers spanning at least one exon junction where possible. For internal exon 10 regions not spanning exon junctions, RNA was treated with ezDNAse (Invitrogen) prior to cDNA preparation.

### Indexing and sequencing

Each sample was dual-indexed via PCR, purified and quantified using Qubit 1× dsDNA HS (Invitrogen). SGE samples were pooled so that gDNA samples were allocated at least 5×10^6^ reads, cDNA samples 3×10^6^ reads, HDR libraries 1×10^6^ reads and negative controls 0.5×10^6^ reads. Purified amplicon pools were sequenced on an Illumina Nextseq or Novaseq according to Illumina protocols, using 300-cycle paired-end runs.

### Processing SGE sequencing data

A published pipeline^12^ was modified to obtain variant counts for each sample from sequencing data. Briefly, samples were demultiplexed using bcl2fastq2 v2.17. SeqPrep was used to trim adapters and merge paired-end reads. Reads were aligned to a reference sequence for each SGE experiment using ‘needleall’ from the EMBOSS v6.6 package. A custom python script was then used to calculate variant frequencies per sample, annotate editing events and add CADD (v1.6) annotations. In order to diminish effects of mutations arising during PCR and sequencing error, reads required one variant from the library and at least one PAM-blocking edit to be included for calculating function scores.

### Deriving HAP1 function scores

Function scores were calculated as the log_2_-ratio of each SNV’s frequency at day 14 to its frequency on day 5. Filters were applied to exclude variants with frequency < 10^−4^ in the HDR library or < 10^−5^ on day 5. Variants for which function scores were discordant between replicates were also filtered; specifically, variants were removed if the difference in function score between replicates was > 1.5, unless both scores were < −1.0. Variants resulting in a different amino acid change in the presence of the PAM-blocking edit were excluded.

Function scores were normalised such that median nonsense and synonymous scores for each coding region matched global median scores. In cases where variants were initially assigned two scores due to SGE regions overlapping, scores from only a single region were ultimately used in analysis, though all overlapping scores are reported in **Table S1**. Scores from the promoter region were used for variants overlapping the 5’-UTR region, and scores from the exon 12-3’ region were used for variants overlapping the exon 12-5’ region.

Function scores for 1- and 5-bp deletions included in the HDR library were calculated and filtered the same way. Where two separate function scores were assigned to the same programmed deletion due to either one or two PAM-blocking edits being present, the function score derived from reads carrying both PAM-blocking edits was used.

### Deriving HAP1 function classes

Statistical testing was used to determine whether function scores were significant. For each coding SGE region, a null distribution was defined using synonymous variants with normal RNA scores (> −1.0). For non-coding regions, the null distribution included all variants. The ‘pnorm’ function in R was used to model the null distribution and calculate p-values, with Benjamini-Hochberg adjustment (‘p.adjust’) to correct for multiple hypothesis testing. A q-value threshold of 0.01 was used to distinguish significantly depleted variants from neutral variants. To categorise depleted variants as ‘LoF’ or ‘intermediate’, we determined function scores significantly different from those of nonsense variants using the distribution of scores for all 155 depleted nonsense variants across SGE regions. We again performed a Benjamini-Hochberg adjustment and set a q-value threshold of 0.01, corresponding to function score threshold of −0.799 to distinguish ‘intermediate’ and ‘LoF’.

### RNA scores

RNA scores were calculated for each coding variant scored in the HAP1 assay as the log2-ratio of each SNV’s cDNA frequency to its gDNA frequency at the day 5 timepoint. RNA scores were normalised such that the median synonymous RNA score equalled 0.0.

### Deriving HMEC function scores and classes

HMEC function scores were calculated for each variant as the mean of the log2-ratio of day 14 frequency to day 7 frequency and the log_2_-ratio of day 21 frequency to day 14 frequency. The same frequency filters were applied as for HAP1. Variants were removed if the difference in function scores between replicates was > 2.0, unless both scores were < −1.0 or both scores were > −1.0. Function scores were normalised between experiments as for HAP1, and the same approach was used to define significantly depleted variants, which were all deemed ‘LoF’. (No intermediate class was defined for HMEC.)

### Deriving combined HAP1–HMEC function classes

Four combined HAP1–HMEC classes were defined based on combinations of function classes in HAP1 and HMEC: ‘concordant-neutral’, ‘HAP1-intermediate, HMEC-neutral’, ‘HAP1-LoF, HMEC-neutral’ and ‘concordant-LoF’. A small number of HMEC-depleted variants scoring neutral (*n* = 9) or intermediate (*n* = 2) in HAP1 were excluded from analyses of combined function classes due to insufficient power. Discordantly scored variants are those that were significantly depleted in HAP1 but not HMEC (i.e., HAP1-LoF, HMEC-neutral and HAP1-intermediate, HMEC-neutral).

### Structural analysis

BRCA1 protein domains were defined as: p.2–101, RING domain; p.1392–1422, CC domain; p.1646–1863, BRCT domain. To visualise CC-domain function scores on the protein structure, the mean function score for missense variants at each amino acid position was calculated and mapped onto the BRCA1–PALB2 CC domain structure (Protein Data Bank (PDB): 7K3S) using PyMOL v2.6.0.

To assess mutations’ effects on BRCA1 stability, FoldX 5.0 (https://foldxsuite.crg.eu/) was used to calculate ΔΔG predictions for all missense substitutions for which SGE function scores were obtained in the RING, CC and BRCT domains. Protein structures were downloaded from the PDB for the BRCA1 RING domain in complex with BARD1 (PDB: 1JM7), the BRCT domain (PDB: 1T15) and the CC domain in complex with PALB2 (mouse structure, PDB: 7K3S). FoldX ‘RepairPDB’ was first used to repair the PDB files, before ‘PositionScan’ was used to calculate ΔΔG predictions. Predictions for three of 28 amino acid positions that differ between the mouse and human CC domain proteins were excluded.

### Predicting transcription factor binding

In the promoter region assayed by SGE, JASPAR predicts a binding site for DP1 (encoded by *TFDP1*). The JASPAR scan tool (https://jaspar.elixir.no/) was used to predict a binding score for each SNV within the 8-bp DP1 binding motif, using matrix profile MA1122.2.

### Comparisons to computational predictions

CADD scores (v1.6) were obtained for all SNVs. Variant consequence labels were determined using CADD annotations for transcript CCDS11456.2. The ‘splice region’ label describes variants immediately adjacent to canonical splice sites that do not alter amino acid sequence, including variants 3–8 bp into introns and variants 1–3 bp into exons.

For missense variants, EVE scores^59^ (https://evemodel.org/; 24 October 2024) and AlphaMissense pathogenicity scores^60^ (https://alphamissense.hegelab.org/; 21 December 2023) were obtained. SpliceAI masked scores were generated by running SpliceAI^24^ with a maximum distance of 500 bp for SNVs within the *BRCA1* transcript (ENST00000357654.9). A ‘maximum SpliceAI score’ for each variant was assigned as the maximum of the ‘acceptor gain’, ‘donor gain’, ‘acceptor loss’ and ‘donor loss’ scores.

### Comparison with other BRCA1 functional assays

Outcomes for missense variants in BRCA1 assays collated by Lyra et al. were obtained^18^. For each variant scored in both the HAP1 and HMEC SGE assays, the proportion of other BRCA1 assays for which the variant was deemed to be functionally abnormal was calculated. Variants were only used for analysis if classifications were provided for at least 3 assays. Prior SGE data^7^ was not considered in this analysis.

### Testing gene essentiality

*BRCA1* essentiality was established in the *BRCA1*^+/−^ HMEC line by analysing indel depletion from day 7 to day 14 after editing the 5’ region of exon 12 according to the HMEC SGE protocol described above. Indel function scores, defined for this experiment as the log_2_-ratio of indel frequency on day 14 over day 7, were determined for the most frequently observed indels at the cut site.

*PALB2* and *RNF168* were edited in exon 4 and exon 1, respectively, via transfection of synthetic gRNAs to the *BRCA1*^+/−^ HMEC line, as previously described. Cells were sampled on days 2 and 14, gDNA was purified, and targeted regions were amplified by PCR and sequenced using NGS. For indels observed at a frequency > 5×10^−4^ on day 2, an indel function score was calculated as the log_2_-ratio of day-14 frequency to day-2 frequency, normalised such that the median score for in-frame indels equalled 0.0. Wilcoxon ranked-sum tests were used to compare scores of frameshifting and in-frame indels.

### Performing SGE in a *RNF168*^−/−^ HMEC line

A synthetic gRNA was used to target coding sequence in exon 1 of *RNF168* and limiting dilution was performed to generate clonal lines from parental *BRCA1*^+/−^ HMECs. The targeted region was Sanger-sequenced and DECODR^61^ was used to deconvolute traces. A line with frameshift mutations on both alleles (c.163_175del and c.166_175del) deemed ‘*RNF168*^−/−^’ was used to perform SGE of exon 12, as described. To allow comparison between the parental line and the *RNF168*^−/−^ line, function scores were normalised so that the median nonsense and median synonymous scores from each line matched.

### ClinVar analysis

ClinVar data was downloaded on 13 September 2024. ClinVar interpretations were simplified for analyses; ‘benign’ and ‘likely benign’ were combined as ‘benign/likely benign’ (B/LB) and ‘pathogenic’ and ‘likely pathogenic’ were combined as ‘pathogenic/likely pathogenic’ (P/LP). VUS and variants with conflicting interpretations of pathogenicity were grouped together as simply ‘VUS’. Receiver operating characteristic (ROC) curves were generated in R using ‘geom_ROC’ from the plotROC package, using B/LB and P/LP variants with at least a one-star review status. For comparison of classification performance across lines, prior HAP1 SGE data for exon 3 was included. For analyses of VUS, all variants were included independent of ClinVar star status.

### Population sequencing data

Allele counts from gnomAD v4.1.0, which includes 416,555 UK Biobank and 314,392 non-UK Biobank exomes, were accessed on 12 February 2025 (https://gnomad.broadinstitute.org/). All of Us data were accessed on 26 June 2024 (https://allofus.nih.gov/). The proportion of variants from each combined function class observed at least once in each population sequencing cohort was determined.

### Estimating breast cancer risk from case–control studies

The BRIDGES dataset from the BCAC used for analysis in this study included *BRCA1* sequencing data from 46,306 breast cancer cases and 43,481 controls. The UK Biobank whole-exome sequencing dataset included a total of 20,553 female breast cancer cases and 227,708 female controls. Dataset quality control and filtering criteria are described elsewhere^47^. Burden analyses in which odds ratios and 95% CIs for breast cancer associated with the presence of any variant in a given SGE-defined function class were estimated using logistic regression. Associations were adjusted for age and study country for the BCAC dataset, and age and genetic ancestry for the UK Biobank dataset. Analyses were performed separately for each dataset and subsequently were combined in a fixed-effects, inverse-variance meta-analysis using the ‘metafor’ R package to derive an overall test of association.

### Assigning ACMG/AMP evidence codes

Truth sets were defined separately for the HAP1 and HMEC SGE datasets by filtering for B/LB and P/LP ClinVar variants with at least a two-star review status and by removing B/LB variants with a maximum SpliceAI score > 0.2. A two-component Gaussian mixture model (GMM) was fitted using Mclust in R and the ‘predict’ function was used to assign a posterior probability of pathogenicity for each variant. Odds of pathogenicity (OddsPath) was calculated as: [*P*2 × (1 − *P*1)] / [(1 − *P*2) × *P*1], where *P*1 and *P*2 are the prior and posterior probabilities of pathogenicity, respectively. To perform this calculation, *P*1 was set uniformly at 0.10, as before^14^. PS3/BS3 evidence codes and corresponding point values^45^ were assigned according to the ACMG/AMP framework^62^ using a maximum evidence strength of ‘strong’, thus capping the points obtainable from a single assay at ±4.

For the HAP1 truth set, the distribution of function scores for B/LB variants was much narrower than the distribution of scores for P/LP variants (**Fig. S7D**). Therefore, a small number of positively scored, neutrally classified variants (function score > 0.35) were not well represented by the GMM. Accordingly, we treated function scores > 0.35 as comprising the tail of the neutral distribution and assigned BS3-strong evidence to match all other HAP1 function scores > −0.19. In both cell lines, evidence codes in favour of pathogenicity were replaced with ‘indeterminate’ for variants not significantly depleted (*q* > 0.01). In HAP1 cells, the strength of evidence in favour of pathogenicity was capped at ‘supporting’ for HAP1-intermediate variants.

For variants assayed in both HAP1 and HMEC, points associated with evidence codes derived from each line were summed to give a combined points value from −8 to +8.

### Statistics

All statistical tests were performed in R v4.3.3 and were two-sided unless otherwise indicated.

## DATA AVAILABILITY

SGE data including all HAP1 and HMEC function scores are available in **Tables S1–S3**. Raw sequencing data (fastq files) from SGE experiments will be made available on the European Nucleotide Archive upon publication (accession: PRJEB95141). BRIDGES summary statistics data and counts are available via the Breast Cancer Association Consortium (BCAC) website, https://www.ccge.medschl.cam.ac.uk/breast-cancer-association-consortium-bcac/data-data-access/summary-results/bridges-summary-results. Individual level data for the BCAC are not publicly available due to ethical review board constraints but are available on request through the BCAC Data Access Co-ordinating Committee. Case-control data from the UK Biobank were accessed under application 102655. Requests for access to UK Biobank should be made to the UK Biobank Access Management Team.

## CODE AVAILABILITY

Custom scripts used to analyse SGE data and to generate figures are available on GitHub: https://github.com/phoebedace/BRCA1_SGE_HAP1_HMEC.

## AUTHOR CONTRIBUTIONS

P.D. led experimental work and analyses. N.F., L.C., M.B., E.M.V, and T.S.W. contributed to experiments. M.Z. and C.T. contributed to analyses. P.D. and G.M.F. wrote the manuscript with input from all authors. G.M.F., K.M., and P.S. supervised the work.

## ACKNOWLEDGEMENTS AND FUNDING

We thank the Crick’s Genomics Scientific Technology Platform (STP) for performing sequencing, the Cell Services STP for maintaining cell lines, the High Throughput Screening STP and Simon Boulton for sharing advice and reagents, and Amanda Spurdle and the ENIGMA consortium for data sharing.

G.M.F. is supported by the Francis Crick Institute, which receives core funding from Cancer Research UK (CC2190), the UK Medical Research Council (CC2190), and the Wellcome Trust (CC2190), and a Cancer Research UK programme award (CG-MAVE). This research has been conducted using the UK Biobank Resource under application 102655. Access to the UK Biobank data has been funded by an internal research grant of the Cyprus Institute of Neurology and Genetics. BCAC is funded by Cancer Research UK (C1287/A16563) and the European Union’s Horizon 2020 Research and Innovation Programme (634935 and 633784 for BRIDGES and B-CAST, respectively). BRIDGES panel sequencing was supported by the European Union’s Horizon 2020 Research and Innovation Program (634935) and the Wellcome Trust (v203477/Z/16/Z). Participating BCAC studies and funders are listed in Zanti et al.^47^

## SUPPLEMENTARY FIGURES

**Supplementary Figure 1.**
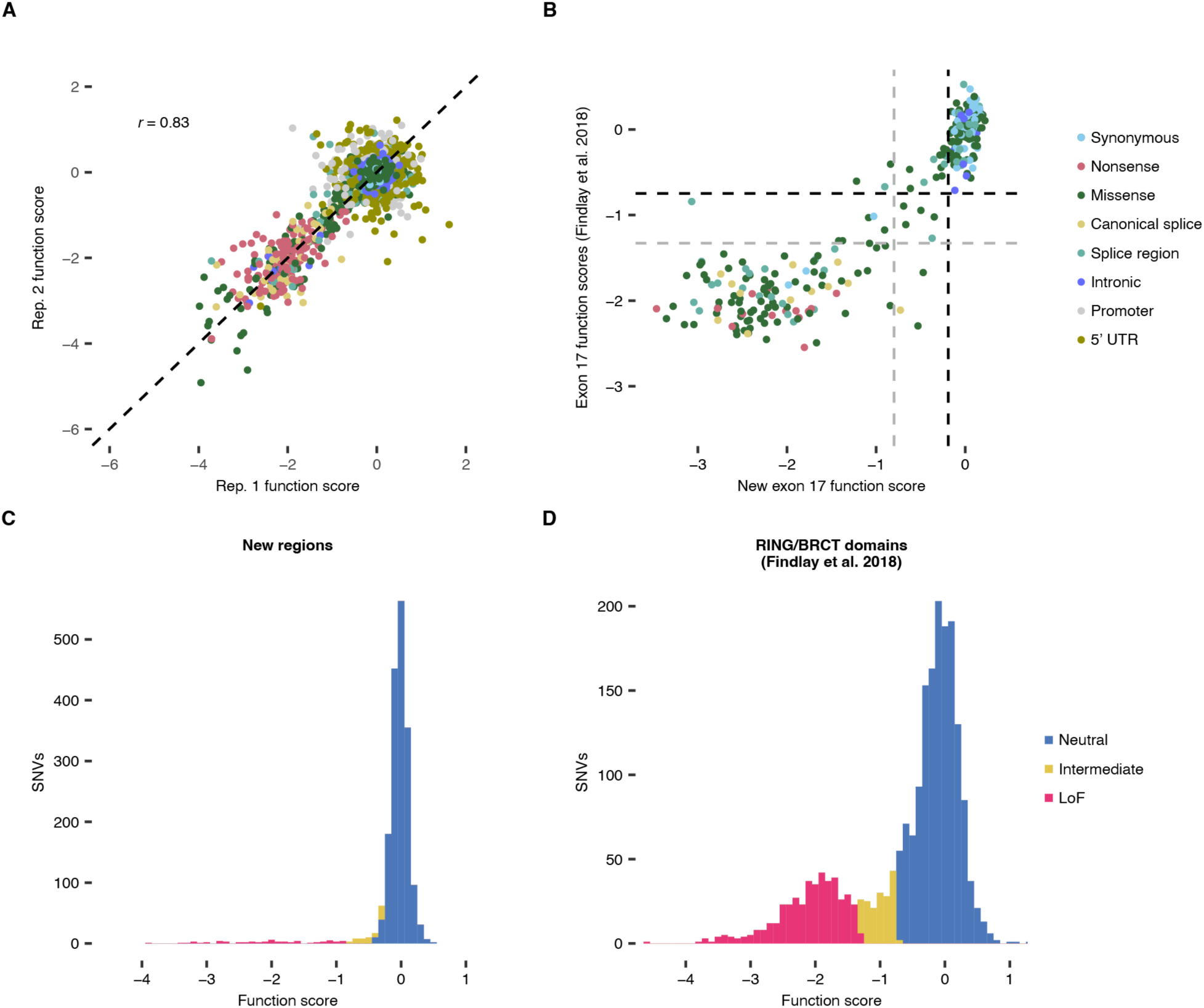
SGE function scores from HAP1 are highly correlated across replicates and with prior SGE data. **A.** The correlation between experimental replicates is shown for function scores of *n* = 4,077 SNVs prior to normalisation and filtering (Pearson’s *r* = 0.83). Only variants outside of the previously assayed RING and BRCT domains are plotted, coloured by mutational consequence. **B.** New function scores for *n* = 287 exon 17 variants are plotted against function scores from Findlay et al.^7^ Black and grey dashed lines divide neutral and intermediate variants, and intermediate and LoF variants, respectively. **C.** Function scores for missense SNVs in newly assayed regions of *BRCA1* (*n* = 1,858). Variants are coloured by function class. **D.** Function scores for missense SNVs in the RING and BRCT domains (*n* = 2,086) assayed previously^7^.

**Supplementary Figure 2.**
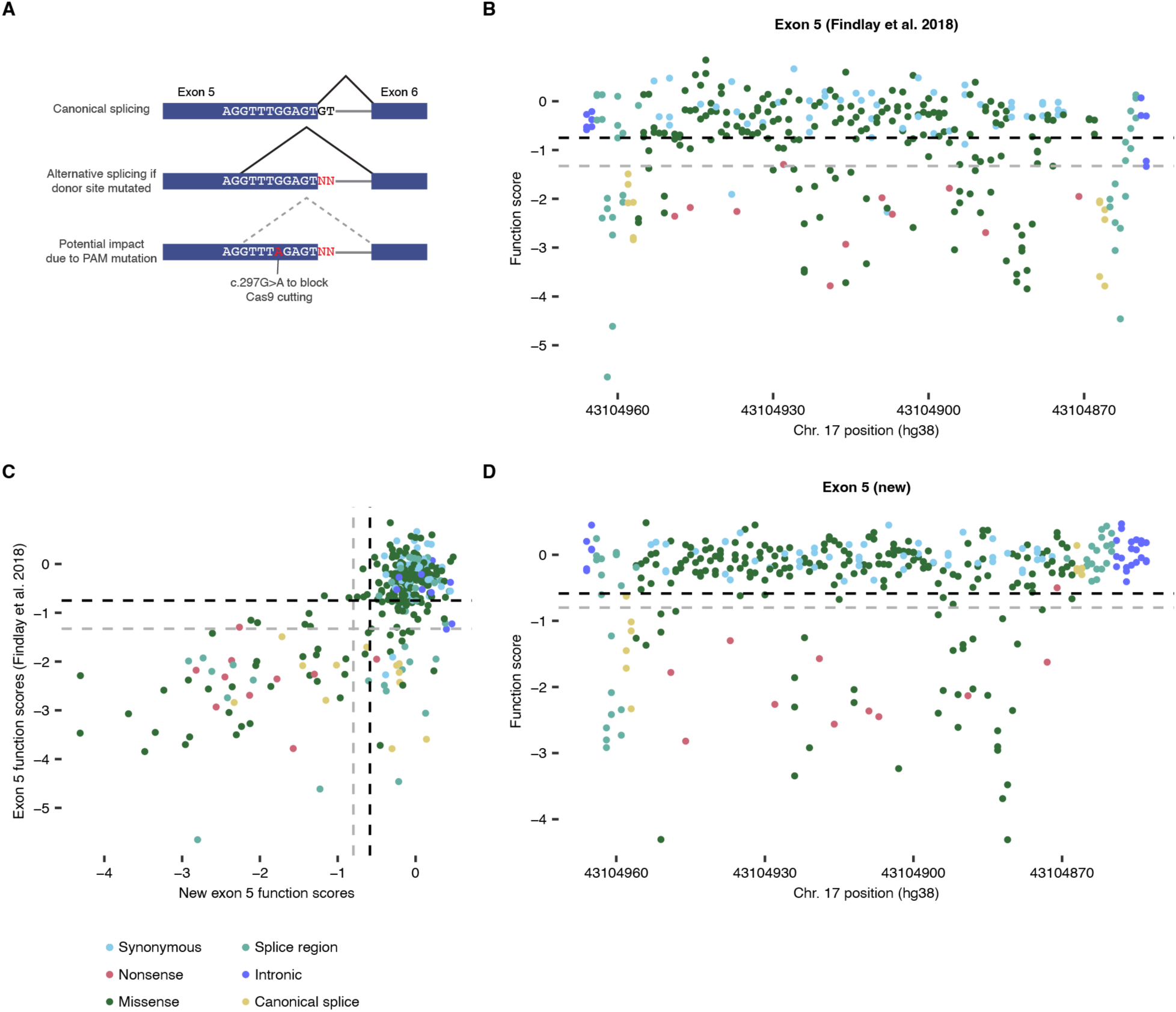
A revised SGE library for exon 5 reveals functional rescue of variants impacting the canonical splice donor site. **A.** Schematic of splicing at the *BRCA1* exon 5 donor site. In the previous SGE experiment from 2018^7^, a synonymous variant, c.297G>A, was introduced to block Cas9 re-cutting following HDR. Splicing analysis^25^ identified an alternative splice site following c.292 potentially capable of rescuing function via an in-frame transcript. As c.297G>A may interfere with alternative splicing, a new exon 5 library lacking c.297G>A was used to perform SGE in this study. **B.** Prior function scores for exon 5, derived using an HDR library containing c.297G>A, are plotted by position. Black and grey dashed lines divide neutral and intermediate variants, and intermediate and LoF variants, respectively. **C.** Prior function scores for exon 5 SNVs are plotted against new function scores derived using an HDR library lacking c.297G>A. **D.** New function scores for exon 5 SNVs are plotted by position.

**Supplementary Figure 3.**
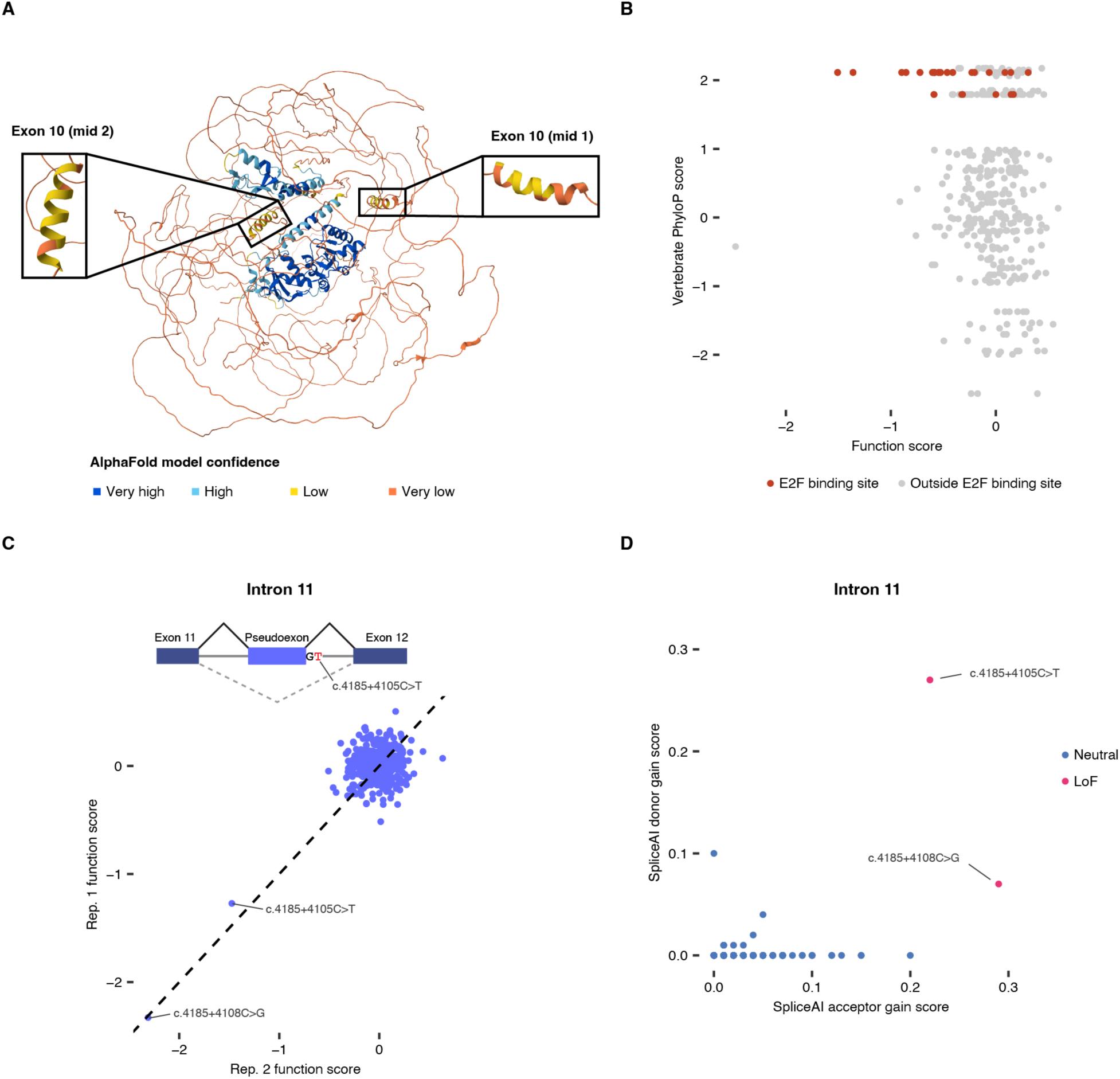
Variants predicted to impact transcription factor binding and splicing lead to functional effects in HAP1. **A.** BRCA1’s protein structure, predicted by AlphaFold2 (AF-P38398-F1-v4), is shown, with residues coloured by model confidence. Internal regions of exon 10 predicted with low confidence to encode alpha helices were assayed in HAP1. Insets have been rotated to show each predicted alpha helix. **B.** HAP1 function scores for SNVs in the promoter and 5’-UTR SGE regions are plotted against vertebrate phyloP scores. SNVs within the predicted 8-bp E2F binding site are coloured red. **C.** The variant c.4185+4105C>T is reported as likely pathogenic and leads to inclusion of a pseudoexon in intron 11^32^. HAP1 function scores are shown per replicate for *n* = 388 intron 11 SNVs assayed. **D.** SpliceAI donor gain and acceptor gain scores are shown for SNVs in the intron 11 region, with variants coloured by function class. A total of 237 variants scored 0.00 for both acceptor gain and donor gain.

**Supplementary Figure 4.**
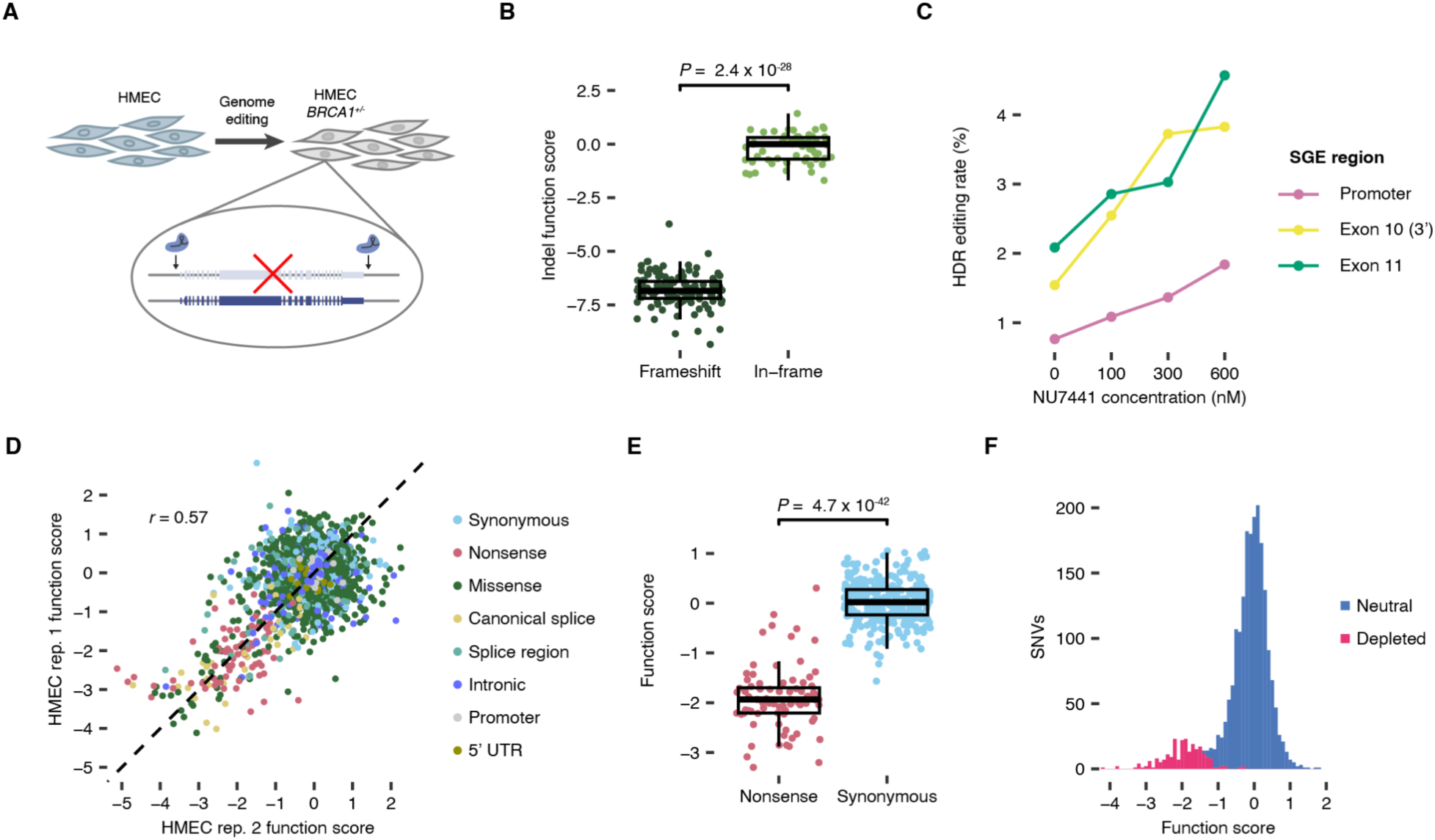
Developing a human mammary epithelial cell line for SGE. **A.** To engineer an HMEC line suitable for SGE, one copy of *BRCA1* was completely deleted by targeting CRISPR to either side of the gene. After isolation of clonal lines, PCR was used to identify lines with whole-gene deletions and to genotype upstream and downstream mutations on the non-deleted allele. **B.** In the *BRCA1*^+/−^ HMEC line selected for SGE, depletion of frameshifting indels (*n* = 120) relative to in-frame indels (*n* = 62) upon targeting of *BRCA1* exon 12 using CRISPR is shown (Wilcoxon Rank-Sum *P* = 2.4×10^−28^). Box plot: centre line, median; box limits, upper and lower quartiles; whiskers, 1.5× interquartile range; all points shown. **C.** The HDR editing rate for three SGE regions is shown for HMECs treated with increasing concentrations of the DNA-PK inhibitor NU7441. **D.** Untreated HMEC function scores for each replicate are plotted for *n* = 2,196 variants, before normalisation and filtering (Pearson’s *r* = 0.57). Variants are coloured by mutational consequence. Scores for exon 17 variants are not included, as final scores were the mean of 4 replicates. **E.** Normalised and filtered function scores from untreated HMECs are shown for *n* = 79 nonsense and *n* = 303 synonymous variants assayed. **F.** The distribution of untreated HMEC function scores coloured by function class is shown.

**Supplementary Figure 5.**
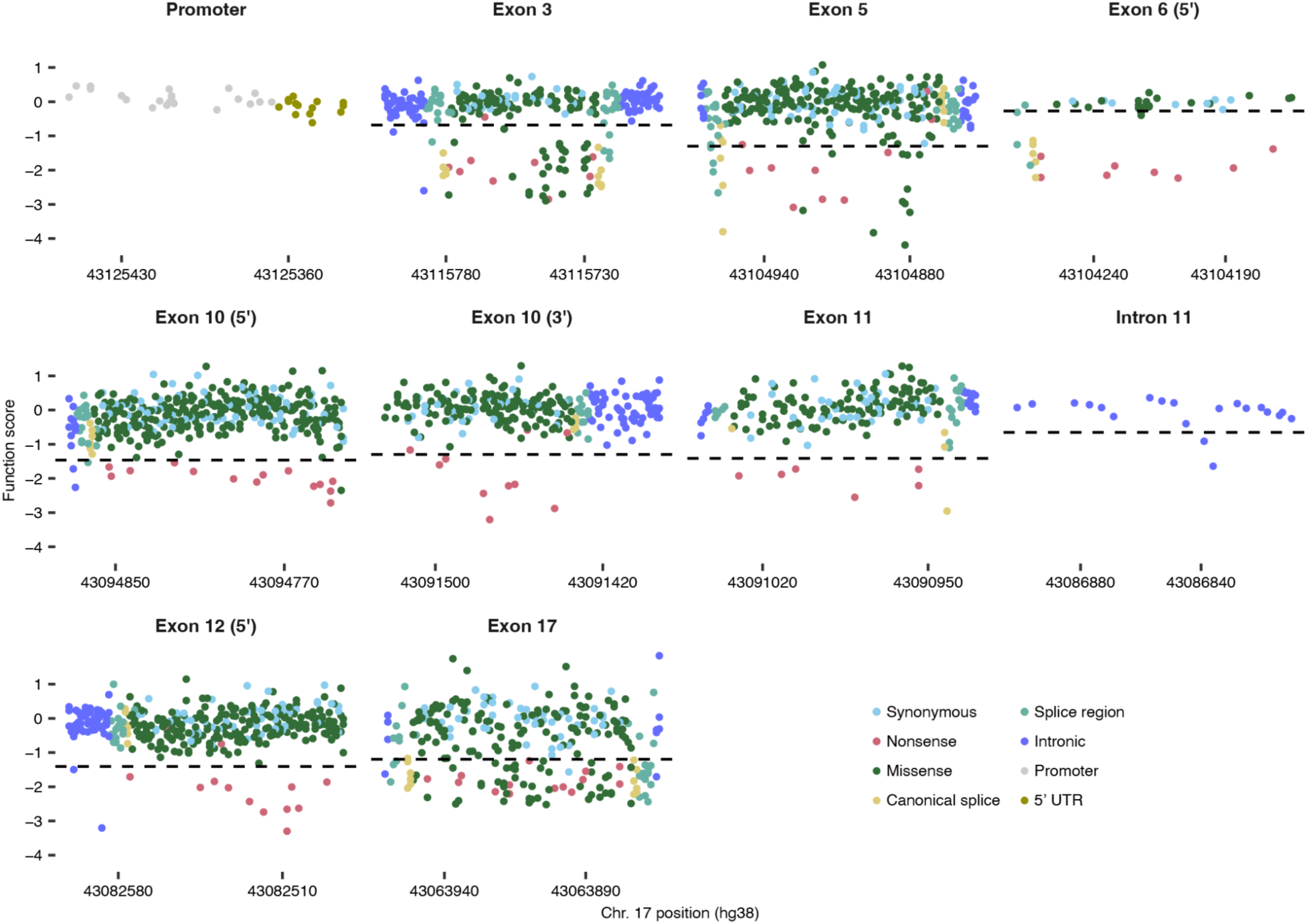
Mapping effects of 2,157 *BRCA1* variants in HMECs. Function scores from HMECs untreated with olaparib are plotted by genomic position for each SGE region. Variants are coloured by mutational consequence. Black dashed lines divide neutral and LoF variants (*q* < 0.01). HDR libraries with a subset of prioritised variants were used for the promoter (*n* = 33), exon 6 (*n* = 48) and intron 11 (*n* = 20) regions due to reduced editing efficiency in HMECs.

**Supplementary Figure 6.**
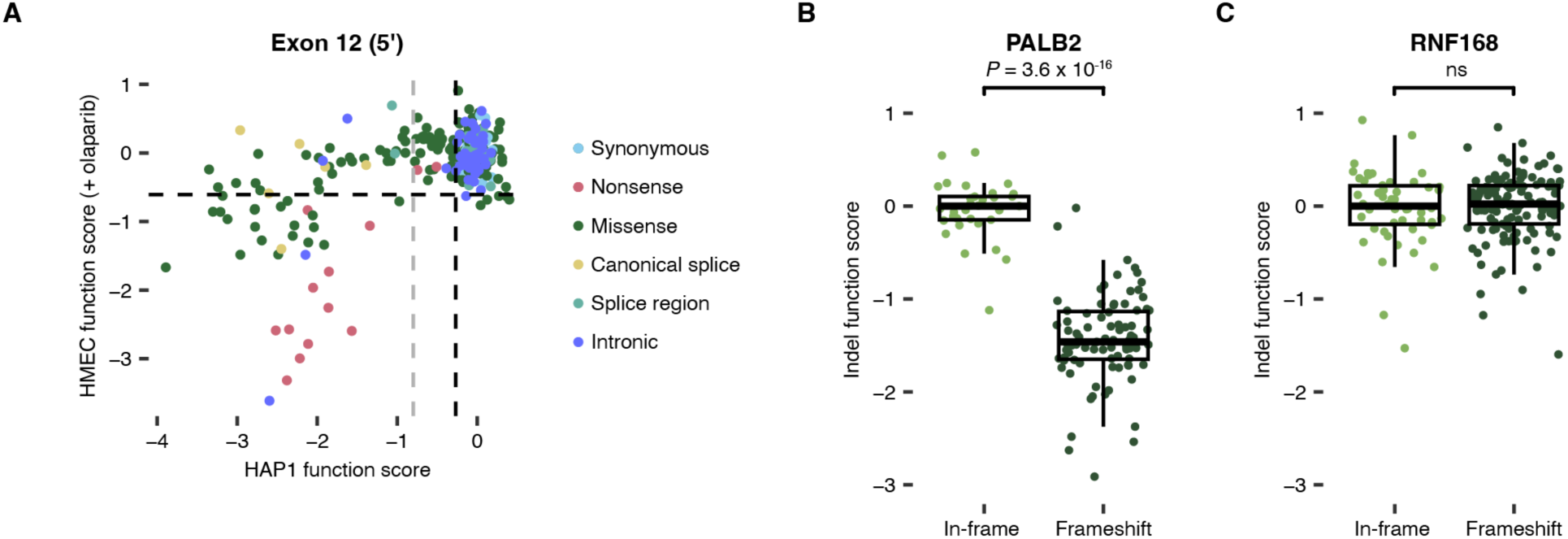
Exploring HMEC sensitivity to functional impairment of the HR pathway. **A.** Function scores for olaparib-treated HMECs are plotted against HAP1-derived function scores for *n* = 275 variants in the 5’ region of exon 12, which encodes the CC domain. Variants are coloured by mutational consequence. Black dashed lines divide neutral and LoF for HMEC and neutral and intermediate for HAP1. The grey dashed line divides intermediate and LoF variants for HAP1. **B,C.** Depletion of frameshifting indels relative to in-frame indels was quantified for *PALB2* (**B**) and *RNF168* (**C**) after targeting each gene using CRISPR. Indel function scores, defined as the log2-ratio of day 14 frequency to day 2 frequency for each indel, reveal selection against frameshifting indels in *PALB2* but not *RNF168* (Wilcoxon Rank-Sum *P* = 3.6×10^−16^ for *PALB2* and *P* = 0.80 for *RNF168*).

**Supplementary Figure 7.**
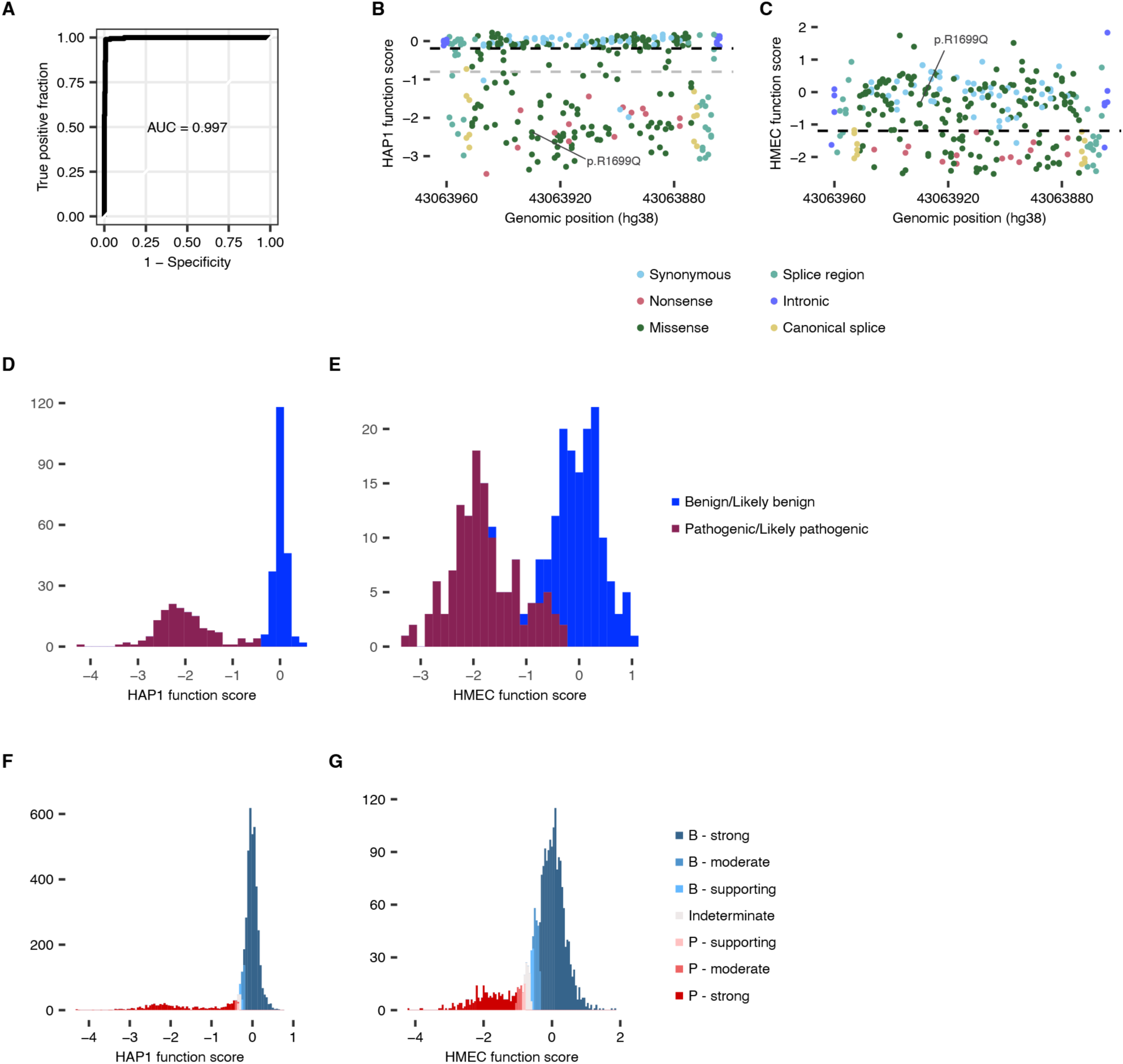
Calibrating function scores to clinical risk and determining evidence codes for variant classification. **A.** ROC curve analysis is shown for discrimination of *n* = 392 B/LB and *n* = 189 P/LP variants in ClinVar using *n* = 4,470 function scores from the complete HAP1 dataset produced in this study. **B,C.** HAP1 (**B**) and HMEC (**C**) function scores for exon 17 are plotted by genomic position and coloured by mutational consequence. In (**B**), the black dashed line divides neutral and intermediate variants, and the grey line divides intermediate and LoF variants. In (**C**), the black dashed line divides neutral and LoF variants. A pathogenic variant of reduced penetrance, p.R1699Q, is labelled in each plot. **D,E.** The distributions of function scores for variants used in truth sets for HAP1 (**D**) and HMEC (**E**) are shown. The HAP1 truth set comprised *n* = 214 B/LB variants and *n* = 152 P/LP variants, while the HMEC truth set comprised *n* = 133 B/LB variants and *n* = 124 P/LP variants, all with two-star status in ClinVar. **F,G.** PS3 and BS3 evidence strengths^45^ were determined for *n* = 4,470 variants assayed in HAP1 (**F**) and *n* = 2,157 variants assayed in HMEC (**G**). The distribution of function scores for each assay is shown, coloured by evidence code assigned prior to any adjustments.

